# Patterns of presentations of children to emergency departments across Europe and the impact of the COVID-19 pandemic: retrospective observational multinational study

**DOI:** 10.1101/2022.03.25.22272926

**Authors:** Ruud G. Nijman, Kate Honeyford, Ruth Farrugia, Katy Rose, Zsolt Bognar, Danilo Buonsenso, Liviana Da Dalt, Tisham De, Ian K. Maconochie, Niccolo Parri, Damian Roland, Tobias Alfven, Camille Aupiais, Michael Barrett, Romain Basmaci, Dorine Borensztajn, Susana Castanhinha, Vasilico Corrine, Sheena Durnin, Paddy Fitzpatrick, Laszlo Fodor, Borja Gomez, Susanne Greber-Platzer, Romain Guedj, Stuart Hartshorn, Florian Hey, Lina Jankauskaite, Daniela Kohlfuerst, Mojca Kolnik, Mark D Lyttle, Patrícia Mação, Maria Inês Mascarenhas, Shrouk Messahel, Esra Akyüz Özkan, Zanda Pučuka, Sofia Reis, Alexis Rybak, Malin Ryd Rinder, Ozlem Teksam, Caner Turan, Valtýr Stefánsson Thors, Roberto Velasco, Silvia Bressan, Henriette A Moll, Rianne Oostenbrink, Luigi Titomanlio, in association with the REPEM network (Research in European Pediatric Emergency Medicine) as part of the EPISODES study group

**Affiliations:** Department of Pediatric Emergency Medicine, Division of Medicine, St. Mary’s hospital - Imperial College NHS Healthcare Trust, London, London, London, UK; Faculty of Medicine, Department of Infectious Diseases, Section of Pediatric Infectious Diseases, Imperial College London, UK; Centre for Pediatrics and Child Health, Imperial College, London, UK; Faculty of Medicine, School of Public Health, Imperial College London, London, UK; Department of Child and Adolescent Health, Mater Dei Hospital, Msida, Malta; Division of Emergency Medicine - Pediatrics, University College London NHS Foundation Trust, London, UK; Department of Pediatric Emergency Medicine, Heim Pal National Pediatric Institute, Budapest, Hungary; Department of Woman and Child Health and Public Health, Fondazione Policlinico Universitario A. Gemelli IRCCS, Rome, Italy; Università Cattolica del Sacro Cuore, Rome, Italy; Division of Pediatric Emergency Medicine, Department of Women’s and Children’s Health, University Hospital of Padova, Italy; Emergency Department & Trauma Center, Ospedale Pediatrico Meyer Firenze, Florence, Italy; SAPPHIRE Group, Health Sciences, Leicester University, Leicester, UK; Pediatric Emergency Medicine Leicester Academic (PEMLA) Group, Leicester Hospitals, Leicester, UK; Pediatric emergency department, Sachs’ Children and Youth Hospital, Stockholm, Sweden; Pediatric Emergency Department, Jean Verdier Hospital, Bondy, France; Pediatric Emergency Department, Children’s Health Ireland at Crumlin, Ireland; Women’s and Children’s Health, School of Medicine, University College Dublin, Dublin, Ireland; National Children’s Research Centre, Crumlin, Dublin, Ireland; Pediatric Emergency Department, Louis Mourier Hospital, Colombes, France; Department of General Pediatrics, Erasmus MC – Sophia, Rotterdam, The Netherlands; Emergency Department, Medisch Centrum Alkmaar, Noordwest Ziekenhuisgroep, Alkmaar, The Netherlands; Hospital Dona Estefania, Centro Hospitalar de Lisboa Central, Portugal; Department of Pediatrics, Paracelsus Medical University, Salzburg, Austria; Pediatric Emergency Department, Children’s Health Ireland at Tallaght, Ireland; Pediatric Emergency Department, Children’s Health Ireland at Temple Street, Ireland; Pediatric Emergency Department, Szent Gyorgy University Teaching Hospital of Fejer County, Szekesfehervar, Hungary; Pediatric emergency department, Cruces University Hospital, Barakaldo, Spain; Biocruces Bizkaia Health Research Institute, Cruces University Hospital, Barakaldo, Spain; Pediatric Emergency Outpatient Clinic, Clinical Division of Pediatric Pulmonology, Allergology and Endocrinology, Department of Pediatrics and Adolescent Medicine, Medical University Vienna, Vienna, Austria; Clinical Division of Pediatric Pulmonology, Allergology and Endocrinology, Department of Pediatrics and Adolescent Medicine, Comprehensive Centre for Pediatrics, Medical University of Vienna, Vienna, Austria; Pediatric Emergency Department, Armand Trousseau Hospital, Paris, France; Pediatric emergency department, Birmingham women’s and children’s NHS Foundation Trust, Birmingham, UK; Birmingham Clinical Trials Unit, Institute of Applied Health Research, University of Birmingham, Birmingham, UK; Pediatric emergency department and pediatric intensive care unit, Dr. von Hauner Children’s Hospital, Ludwig-Maximilians-University Munich, Munich, Germany; Hospital of Lithuanian University of Health Sciences Kauno Klinikos, Lithuania; Department of General Pediatrics, Medical University of Graz, Austria; University Medical Centre Ljubljana, Univerzitetni Klinični Center, Department of Infectious Diseases, Ljubljana, Slovenia; Emergency Department, Bristol Royal Hospital for Children, Bristol, UK; Faculty of Health and Applied Sciences, University of the West of England, Bristol, UK; Pediatric Emergency Service, Hospital Pediátrico, Centro Hospitalar e Universitário de Coimbra, Portugal; Departamento da Criança e do Jovem-Urgencia Pediatrica, Hospital Prof. Doutor Fernando da Fonseca, Amadora, Portugal; Pediatric emergency department, Alder Hey Children’s NHS Foundation Trust, Liverpool, UK; Pediatric Emergency Department, Ondokuz Mayıs University, Samsun, Turkey; Pediatric emergency department, Children’s Clinical University Hospital, Riga Stradins University, Riga, Latvia; Pediatric Department, Centro Hospitalar Tondela-Viseu, Viseu, Portugal; Pediatric Emergency Department, Hopital Universitaire Robert-Debre, Paris, France; ACTIV, Association Clinique et Thérapeutique Infantile du Val-de-Marne, Créteil, France; INSERM, ECEVE, Université de Paris, Paris, France; Pediatric emergency department, Astrid Lindgrens Children’s hospital, Karolinska University, Sweden; Division of Pediatric Emergency Medicine, Department of Pediatrics, Hacettepe University School of Medicine, Ankara, Turkey; Department of Pediatrics, Division of Emergency Medicine, Mersin City Training and Research Hospital, Toroslar, Mersin, Turkey; Childreńs Hospital, Barnaspitali Hringsins, Reykjavik, Iceland; Pediatric emergency unit, Hospital Universitario Río Hortega, Valladolid, Spain

**Author notes:** Corresponding author: (RN). Membership of the EPISODES study group is provided as supporting information.

## Abstract

**Background:** To investigate the impact of the COVID-19 pandemic and infection prevention measures on children visiting emergency departments across Europe.

**Methods:** Routine health data were extracted retrospectively from electronic patient records of children aged <16 years, presenting to 38 emergency departments (ED) in 16 European countries for the period January 2018 – May 2020, using predefined and standardized data domains. Observed and predicted numbers of ED attendances were calculated for the period February 2020 to May 2020. Poisson models and incidence rate ratios (IRR) were used to compare age groups, diagnoses and outcomes.

**Findings:** Reductions in pediatric ED attendances, hospital admissions and high triage urgencies were seen in all participating sites. ED attendances were relatively higher in countries with lower SARS-CoV-2 prevalence (incidence rate ratio (IRR) 2·62, 95% CI 2·19 to 3·13) and in children aged >12 months (12-<24 months IRR 0·89, 95% CI 0·86 to 0·92; 2-<5years IRR 0·84, 95% CI 0·82 to 0·87; 5-<12 years IRR 0·74, 95% CI 0·72 to 0·76; 12-<16 years IRR 0·74, 95% CI 0·71 to 0·77; vs. age <12 months as reference group). The impact on pediatric intensive care admissions (IRR 1·30, 95% CI 1·16 to 1·45) was not as great as the impact on general admissions. Lower triage urgencies were reduced more than higher triage urgencies (urgent triage IRR 1·10, 95% CI 1·08 to 1·12; emergent and very urgent triage IRR 1·53, 95% CI 1·49 to 1·57; vs. non-urgent triage category). Reductions were highest and sustained throughout the study period for children with communicable infectious diseases.

**Interpretation:** Reductions in ED attendances were seen across Europe during the first COVID-19 lockdown period. More severely ill children continued to attend hospital more frequently compared to those with minor injuries and illnesses, although absolute numbers fell.

**Funding:** RGN was supported by National Institute of Health Research, award number ACL-2018-021-007.

**Trial registry:** ISRCTN91495258

## Introduction

Healthcare systems across Europe continue to be greatly affected by the COVID-19 pandemic. During the initial phase of the pandemic, evidence emerged that children were less likely to develop symptoms of severe acute SARS-CoV-2 infection, when compared with adults. [1–5] In fact, reduced numbers of children visiting urgent and emergency care services were reported following the introduction of infection prevention measures. These seemed to be greatest for infectious communicable diseases. [6–10] Typically, these studies did not compare patterns between countries or in relation to different public health strategies.

At the same time, concerns were raised about potential delays in, and higher acuity of, presentations to appropriate healthcare services, as a result of difficulties accessing these services, changes in health care provision preferencing virtual consultations, fear of exposure to SARS-CoV-2 in health care facilities and blanket ‘Stay at Home’ statements. [11–13] In the UK this resulted in a statement from the Royal Society of Pediatrics and Child Health to reassure parents and caregivers, urging them to seek appropriate urgent and emergency medical attention when worried about the acute illness or injury of their child. [14] Additionally, mostly anecdotal evidence reported increased numbers of specific childhood diagnoses, such as diabetic ketoacidosis [15] and intussusception [16]. These implicated a possible link with SARS-CoV-2 infection, yet evidence from large scale cohorts is lacking. Concerns were also raised for the mental health of children resulting from school closures and stay at home orders. [17,18]

In this study, we aimed to compare the number of children presenting to emergency departments (EDs) across Europe during the first phase of the COVID-19 pandemic with the two previous years; investigating any change in severity of illness and describing the associations with specific diagnoses potentially related to SARS-CoV-2.

## Methods

### Study design, setting and participants

This retrospective, observational study included 38 sites from 16 European countries (S1 Table) as part of the ‘Epidemiology, severity and outcomes of children presenting to emergency departments across Europe during the SARS-CoV-2 pandemic’ (EPISODES) – study (trial registration number: ISRCTN91495258). Sites were selected from the Research in European Pediatric Medicine (REPEM) and the Pediatric Emergency Research in the United Kingdom and Ireland (PERUKI) networks following the earlier work of Bressan et al. [19] Routine clinical data from all children presenting to the ED were extracted from electronic health records for the period January 1^st^ 2018 – May 17^th^ 2020. Upper age limit varied between sites at between 16 and <18 years old.

Aggregated, standardized data were uploaded using the REDCap online platform (S2 File). For the period January 1^st^ 2018 and February 1^st^ 2020 data were collected on a monthly basis. For the period February 2^nd^ 2020 – May 17^th^ 2020 on a weekly basis. This amounted to a total of forty time-windows (S3 Table). The clinical report form included ten different data domains: 1) moment of presentation, 2) patient characteristics, 3) mode of arrival and referral pathway, 4) triage urgency, 5) type of presenting problem, 5) vital signs, 6) diagnostics performed in the ED, 7) treatment in the ED, 8) diagnosis, 9) hospital admission, 10) duration of ED and hospital stay (S2 File); data availability varied between sites (S4 Fig).

Triage urgency levels, used to determine the urgency of care in the ED, were categorized in three categories, defined as emergent-very urgent (or RED-ORANGE, or level 1-2), urgent (or YELLOW, or level 3) and standard-non urgent (or GREEN-BLUE, or level 4-5) to allow uniform coding between sites. For diagnosis coding, ICD-10 codes were issued for guidance (S5 Table), but an internally and temporally consistent coding approach was encouraged for each of the individual sites, acknowledging different coding systems and strategies in the ED. This was checked by plotting the diagnoses coding in time as percentage of total number of attendances for each site. Severity was defined based on level 1-2 urgency classification at triage, any hospital admissions, pediatric intensive care unit (PICU) admission or death in ED.

### Data analyses

The completeness, quality and internal consistency of data were checked by plotting the absolute numbers, as well as percentage of total attendances, for each variable of interest in time for the whole study period 2018 – 2020. In order to quantify changes in attendances we compared observed attendances with predicted numbers of attendances. Predicted numbers of attendances were estimated using monthly data for the 25 months prior to February 3^rd^ 2020. As the data had both a trend and seasonal component we used Holt-Winters exponential smoothing to make short-term monthly forecasts for February, March, April and May 2020. We adjusted these to weekly estimates of predicted numbers. We plotted predicted ED attendances against the introduction of national infection prevention measures (Fig 1, S6 Table). [20] We also calculated twenty-eight day mean numbers for selected diagnoses, PICU and hospital admission, and death in ED for each month from January through to April for the years 2018 to 2020.

**Fig 1.**
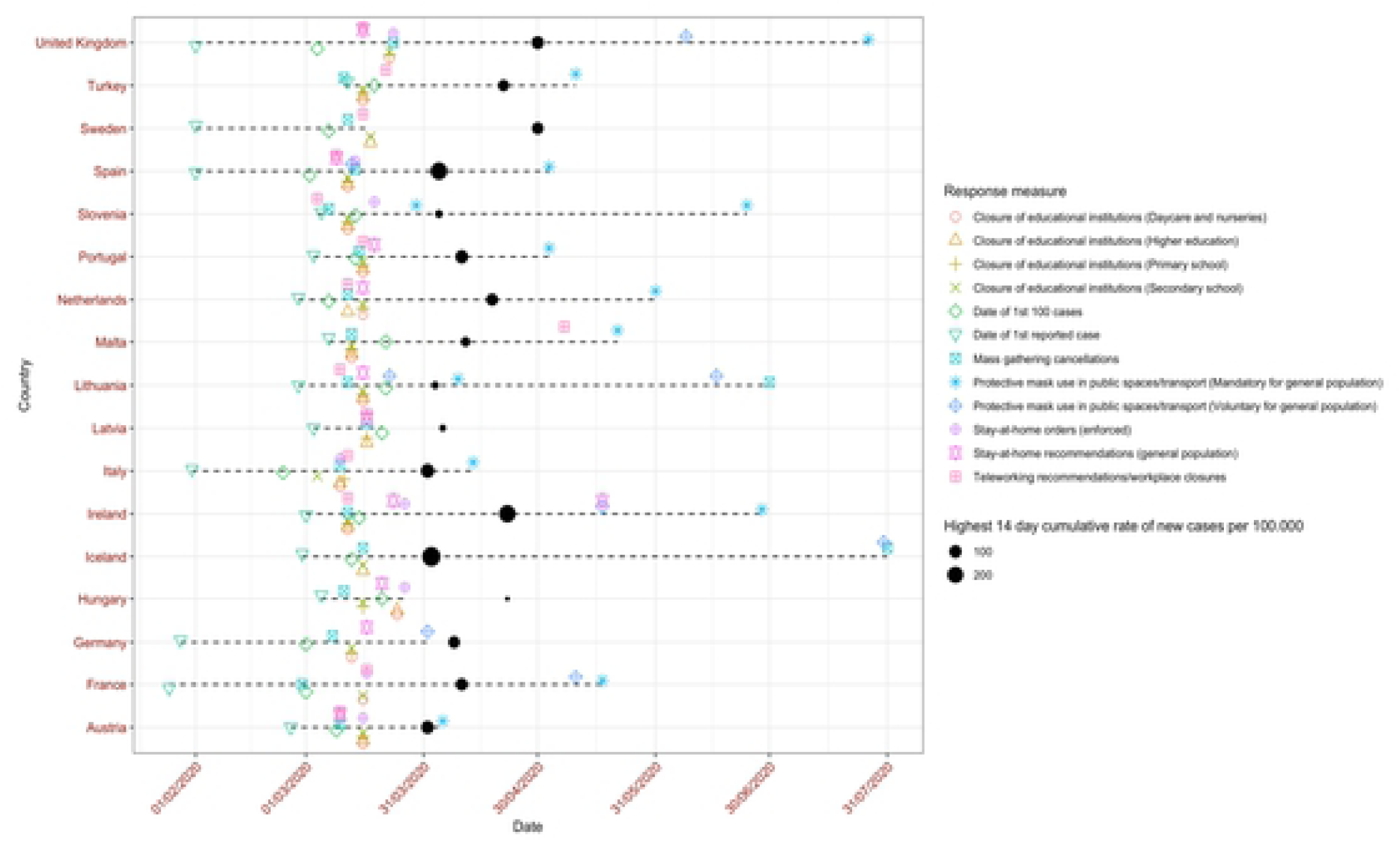
Timelines of first phase of COVID-19 pandemic in participating countries. Timelines of the introduction of national infection prevention measures (‘Response measures’), as well as dates for the first and 1^st^ 100 cases of SARS-CoV-2 for each of the countries participating in the EPISODES study. The black circle depicts the date of the highest 14 day cumulative rate of new SARS-CoV-2 cases per 100,000, with the size reflecting the actual case rate.

We used a Poisson model, adjusted for time since February 3^rd^ 2020, to determine if there were differences between age groups, diagnosesand disposition for patients. For each model, the outcome was the count of attendances per week from the week beginning February 3^rd^ 2020 to the week beginning May 4^th^ 2020, with an offset of the predicted number of attendances in each week. For age groups, the analysis was adjusted for site; for diagnoses and disposition, numbers were too small to make forecasts at site leveland we therefore aggregated these across the whole sample. For diagnoses, we completed two models, one with eight separate diagnoses and one where these were divided into three groups: surgical presentation (ie. appendicitis), communicable diseases (ie. tonsillitis, otitis media, lower respiratory tract infection (LRTI) and gastroenteritis)and ‘other’ (ie. mental health issue, radius fracture and minor head trauma). For three diagnosis groups the number of attendances were too low to make sensible forecasts, namely diabetic keto-acidosis, testicular torsionand the combined group of intussusception, volvulusand malrotation. In addition, we determined if there were associations between the change in hospital attendances and the prevalence of SARS-CoV-2 in the country, as per the European Centre for Disease Prevention and Control (ECDC) (Fig 1, S7 Table)and the number of COVID-19 measures that were introduced in each hospital in response to the pandemic as previously detailed by Rose et al. [21] Rose et al. performed a survey study describing all major changes in local and regional health care pathways, including the diverting of patient groups to or away from the ED, and service provision in each of the study sites. High prevalence countries were defined as a cumulative 14-day rate of >80 new cases per 100,000 of the population. For countries where there was more than one site per country we used an ANOVA to determine if there was evidence that within country differences were greater than between country differences, for total attendances in March and April, adjusted for predicted numbers to account for differences in site sizes. One site (MAL001) was unable to provide information on diagnosis so it was excluded from this section of analysis; three sites (SLO001, POR005, TUR001) did not provide triage data. Two sites were excluded (NL002 and HUN002) from the forecasting analyses and Poisson models as they had missing data in the period before the pandemic (2018). One site (IRE003) was excluded from the Poisson models because it closed to pediatric attendances in response to the pandemic. One site (TUR003) accounted for 18% of all attendances and we carried out sensitivity analysis to confirm the impact of this site on all results. Analyses were performed using R v4.0.0.

### Ethics

Following initial approval by the UK Health Research Authority, all participating sites obtained approval from their national/local institutional review boards (S8 Table). The need for individual patient informed consent was waived. Data sharing agreements were in place.

## Results

### Description of sites

Sites included in the study varied in terms of size and service provision (S1 Table). The annual number of ED attendances ranged from 4,961 (NL001, 2019) to 295,787 (TUR003, 2019) (S9 Fig). All but three sites were tertiary academic hospitals with specialized pediatric emergency departments; the remaining three sites were general teaching hospitals, two of which had dedicated pediatric sections and staff, and one of which had a mixed emergency department. Sites in Austria, Sloveniaand Netherlands mainly saw medical presentations, whereas the other sites saw both medical and surgical/trauma presentations.

### Description of infection prevention measures and SARS-CoV-2 prevalence

Timing and degree of infection prevention measures were similar across European countries (Fig 1; S6 Table). Notably, Iceland and Sweden did not close day-care, nurseries, or primary education; Germany and the United Kingdom kept higher education open; Sweden did not close any public spaces; Hungary and Sweden did not advocate use of face masks; Malta, Icelandand Sweden did not introduce stay-at-home recommendations; and Germany, Hungaryand Iceland did not formally close workspaces. Highest national prevalence of SARS-CoV-2 varied between countries (Fig 1, S7 Table).

### Impact on total attendances

All 38 sites had significant reductions in attendances in spring 2020 (Table 1, Fig 2). The largest reduction was seen in AUS001 with observed numbers at 5% (95% CI 5% to 6%) of predicted in the week starting 30/03/2020; the smallest peak reduction in ED attendances was at 56% (96% CI 52% to 60%) of predicted in SWE001 during the same week. IRE003 closed for pediatric visits from 30/03/2020 onwards, with most patients diverted to IRE001. Poisson models, adjusted for time since intervention and predicted numbers of attendances showed that there were significant differences between sites.

**Fig 2.**
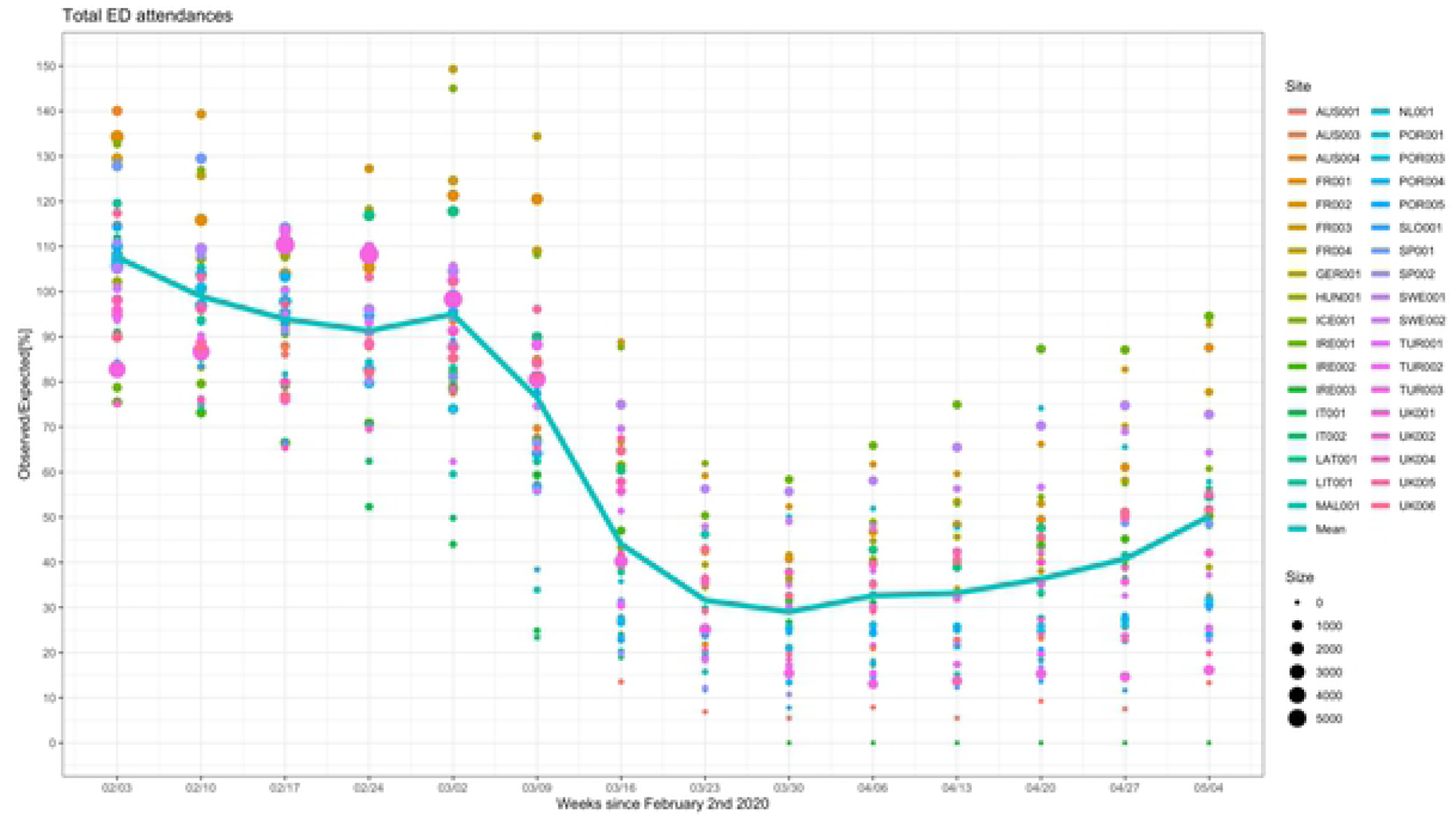
Observed and predicted in % for the number of total emergency department attendances. The observed and predicted number of children presenting to emergency departments in countries across Europe in the weeks following February 2^nd^ 2020 until May 11^th^ 2020, for all sites combined. The colour and the size of the dots reflect the actual number of ED attendances for each site and for each time window. The line connects the mean of the observed vs predicted point estimates for each of the individual sites for each time window.

**Table 1.**
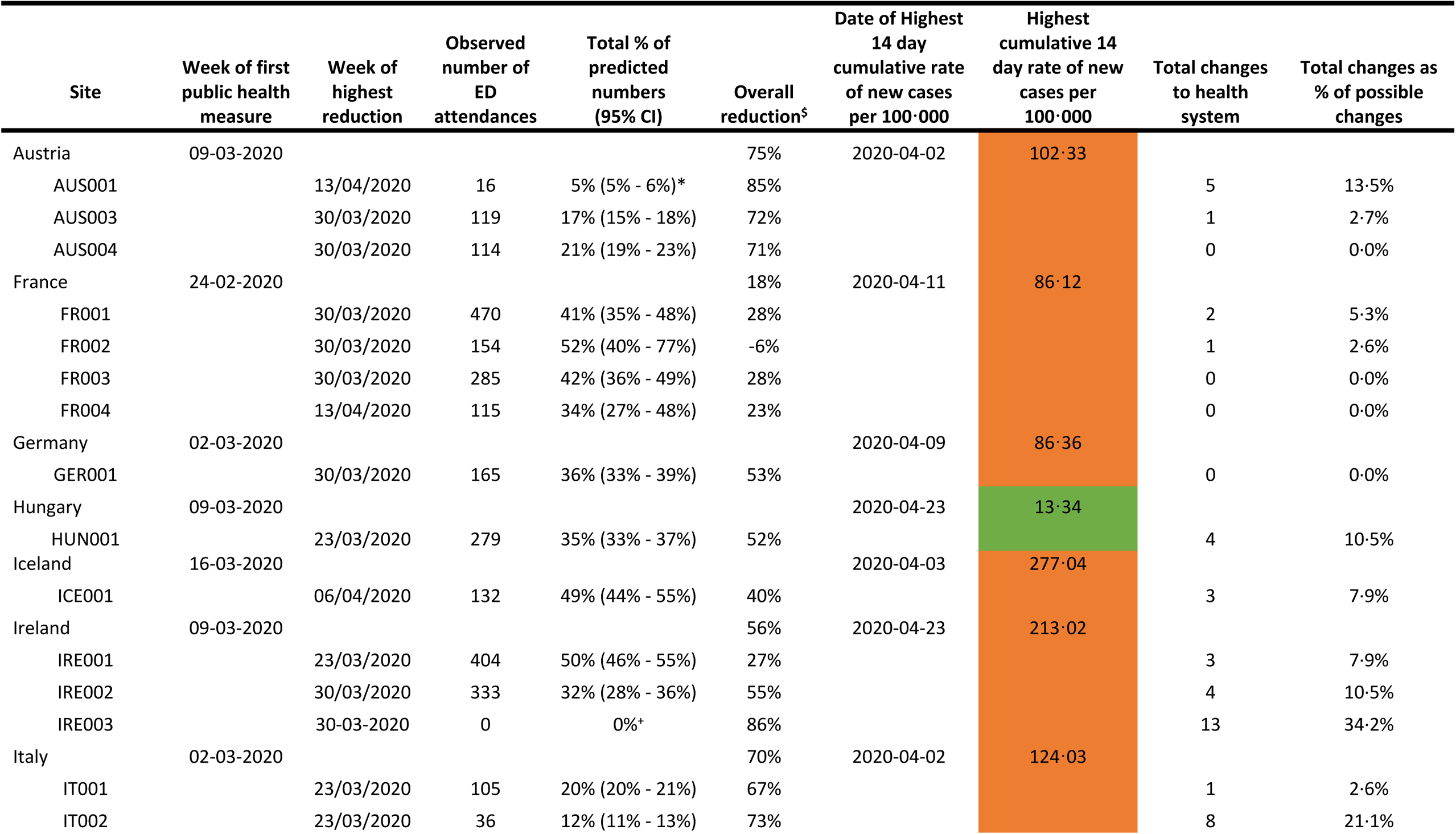
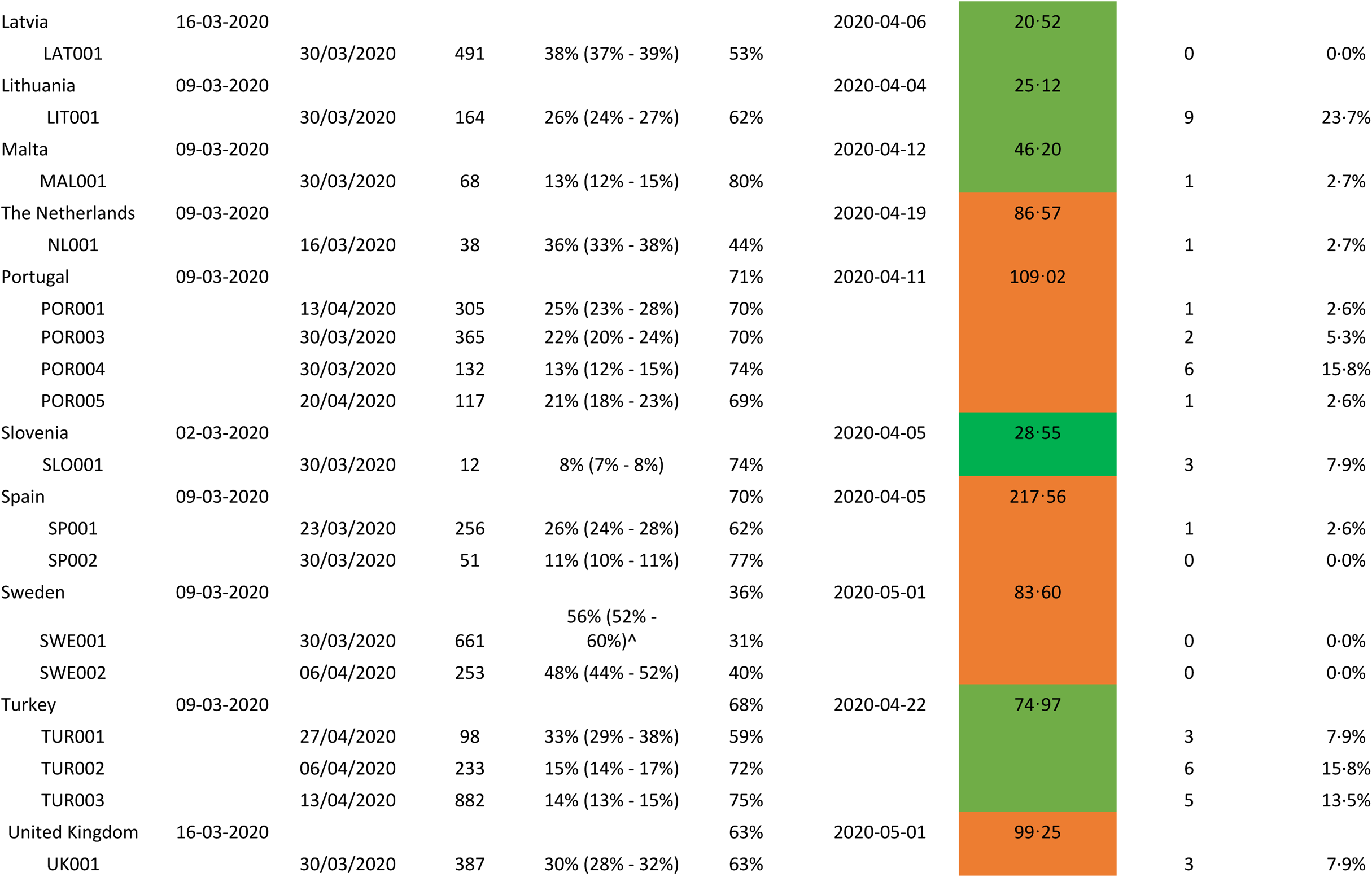
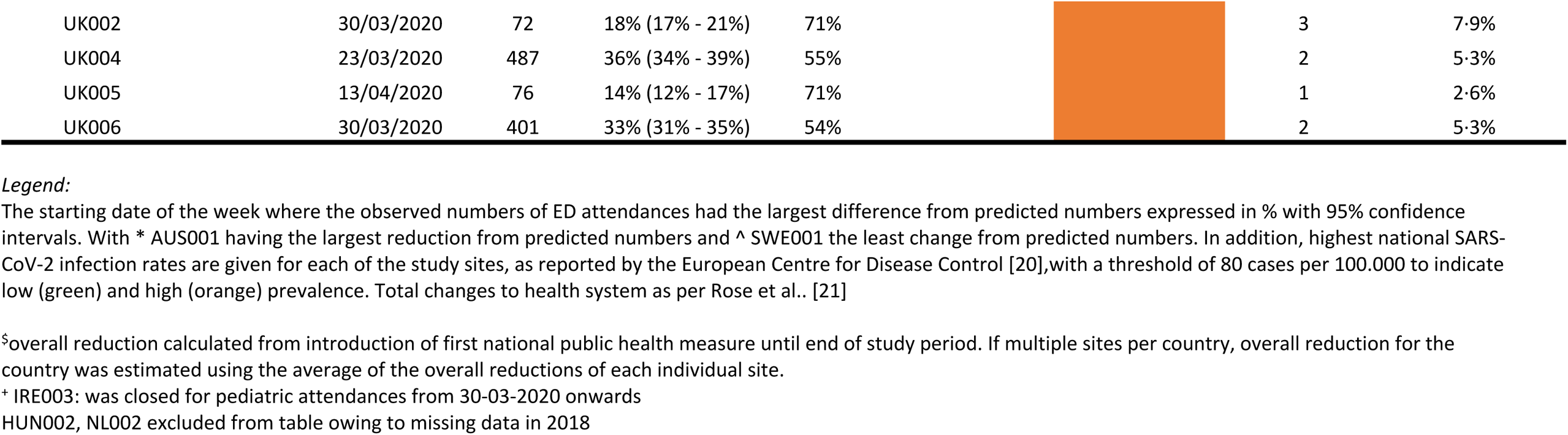
Summary data on the lowest observed number of ED attendances during COVID-19 for each participating centre. The starting date of the week where the observed numbers of ED attendances had the largest difference from predicted numbers expressed in % with 95% confidence intervals. With * AUS001 having the largest reduction from predicted numbers and ^ SWE001 the least change from predicted numbers. In addition, highest national SARS-CoV-2 infection rates are given for each of the study sites, as reported by the European Centre for Disease Control [20],with a threshold of 80 cases per 100.000 to indicate low (green) and high (orange) prevalence. Total changes to health system as per Rose et al.. [21]

Observed attendances, with respect to predicted, were relatively higher in sites in France, Sweden, Ireland, Iceland, Latviaand the Netherlands, where observed attendance rates were greater than 50% of predicted. However, there was considerable overlap between all sites when 95% confidence intervals were considered. Results of the Poisson models suggest that attendances in Spring 2020 were higher in EDs in countries with lower SARS-CoV-2 prevalence (incidence rate ratio (IRR) 2·62, 95% CI 2·19 to 3·13) (Table 2). We found a relationship between the number of introduced organisational COVID-19 measures and ED attendancesand more organisational COVID-19 measures were associated with lower numbers of ED attendances when adjusted for predicted ED attendances (IRR 0·13, 95% CI 0·11 to 0·16, when sites with four or more measures were compared to sites with no measures). ED attendances across all age groups significantly reduced (S10 and S11 Figs). Attendances in children aged above 12 months were reduced more than children below 12 months (12-<24 months IRR 0·89, 95% CI 0·86 to 0·92; 2-<5years IRR 0·84, 95% CI 0·82 to 0·87; 5-<12 years IRR 0·74, 95% CI 0·72 to 0·76; 12-<16 years IRR 0·74, 95% CI 0·71 to 0·77; vs. age <12 months as reference group) (Table 2). There was insufficient evidence to conclude that this pattern continued with increasing age for children aged 12 months and older. Patterns between sites within the same country appeared similar (S12 Fig) with strong evidence that between country differences were greater than within country differences (F value:6·453; p:0·002).

**Table 2.** Poisson regression models for ED attendances

|  | Incidence Rate Ratio<br>(95% Confidence Interval) | *p value* |
| --- | --- | --- |
| **Number of COVID-19 measures in hospital<sup>§</sup>** |  |  |
| No measures – reference group |  |  |
| 1 measure | 0.426 (0.391 to 0.463) | <0.0001 |
| 2 measures | 0.760 (0.717 to 0.806) | <0.0001 |
| 3 measures | 0.454 (0.413 to 0.499) | <0.0001 |
| 4+ measures | 0.130 (0.108 to 0.155) | <0.0001 |
| **SARS-CoV-2 prevalence<sup>^</sup>** |  |  |
| Low prevalence | 2.616 (2.197 to 3.144) | <0.0001 |
| **Age Group** |  |  |
| 0-<12 months – reference group |  |  |
| 12-<24 months | 0.888 (0.859 to 0.917) | <0.0001 |
| 2-<5 years | 0.841 (0.816 to 0.866) | <0.0001 |
| 5-<12 years | 0.736 (0.712 to 0.761) | <0.0001 |
| 12-16 years | 0.744 (0.714 to 0.773) | <0.0001 |
| **Triage urgency classification** |  |  |
| Non-urgent and standard triage categories – reference group |  |  |
| Urgent | 1.096 (1.076 to 1.116) | <0.0001 |
| Emergent and very urgent | 1.530 (1.488 to 1.573) | <0.0001 |
| **Diagnosis I** |  |  |
| Appendicitis – reference group |  |  |
| Gastro-intestinal infections | 0.279 (0.253 to 0.308) | <0.0001 |
| Minor head injury | 0.783 (0.709 to 0.866) | <0.0001 |
| LRTI | 0.357 (0.323 to 0.396) | <0.0001 |
| Mental health issues | 0.688 (0.609 to 0.777) | <0.0001 |
| Otitis media | 0.231 (0.206 to 0.260) | <0.0001 |
| Radius fracture | 0.732 (0.654 to 0.819) | <0.0001 |
| Tonsillitis | 0.189 (0.172 to 0.208) | <0.0001 |
| **Diagnosis II** |  |  |
| Surgical presentation – appendicitis – reference group |  |  |
| Communicable diseases | 0.238 (0.253 to 0.308) | <0.0001 |
| Other | 0.754 (0.709 to 0.866) | <0.0001 |
| **Outcome** |  |  |
| Admission – reference group |  |  |
| Death | 1.749 (0.876 to 3.069) | 0.077 |
| PICU Admission | 1.295 (1.157 to 1.445) | <0.0001 |

<sup>§</sup> The number of changes made in each hospital in response to the pandemic as previously detailed by Rose et al.[21]
<sup>^</sup> Low prevalence countries were defined as a cumulative 14-day rate of ≤ 80 new cases per 100,000 of the population as per the European Centre for Disease Prevention and Control (ECDC) [20]
LRTI: lower respiratory tract infection, PICU: pediatric intensive care unit

### Triage urgency

Overall, there was a higher impact (observed compared to predicted) in children with lower triage urgency when compared to children with high triage classification (urgent triage, IRR 1·10, 95% CI 1·08 to 1·12; emergent and very urgent triage IRR 1·53, 95% CI 1·49 to 1·57; vs. non-urgent triage category), even though clear reductions were seen for all triage categories (S13 and S14 Figs).

### Hospital and PICU admissions

Hospital and PICU admissions were fewer than predicted (Figs 3 and 4, S15 Fig). We did not observe an increase in the number of deaths in ED. The impact on PICU admissions (IRR 1·30, 95% CI 1·16 to 1·45) was not as great as the impact on general admissions.

**Fig 3.**
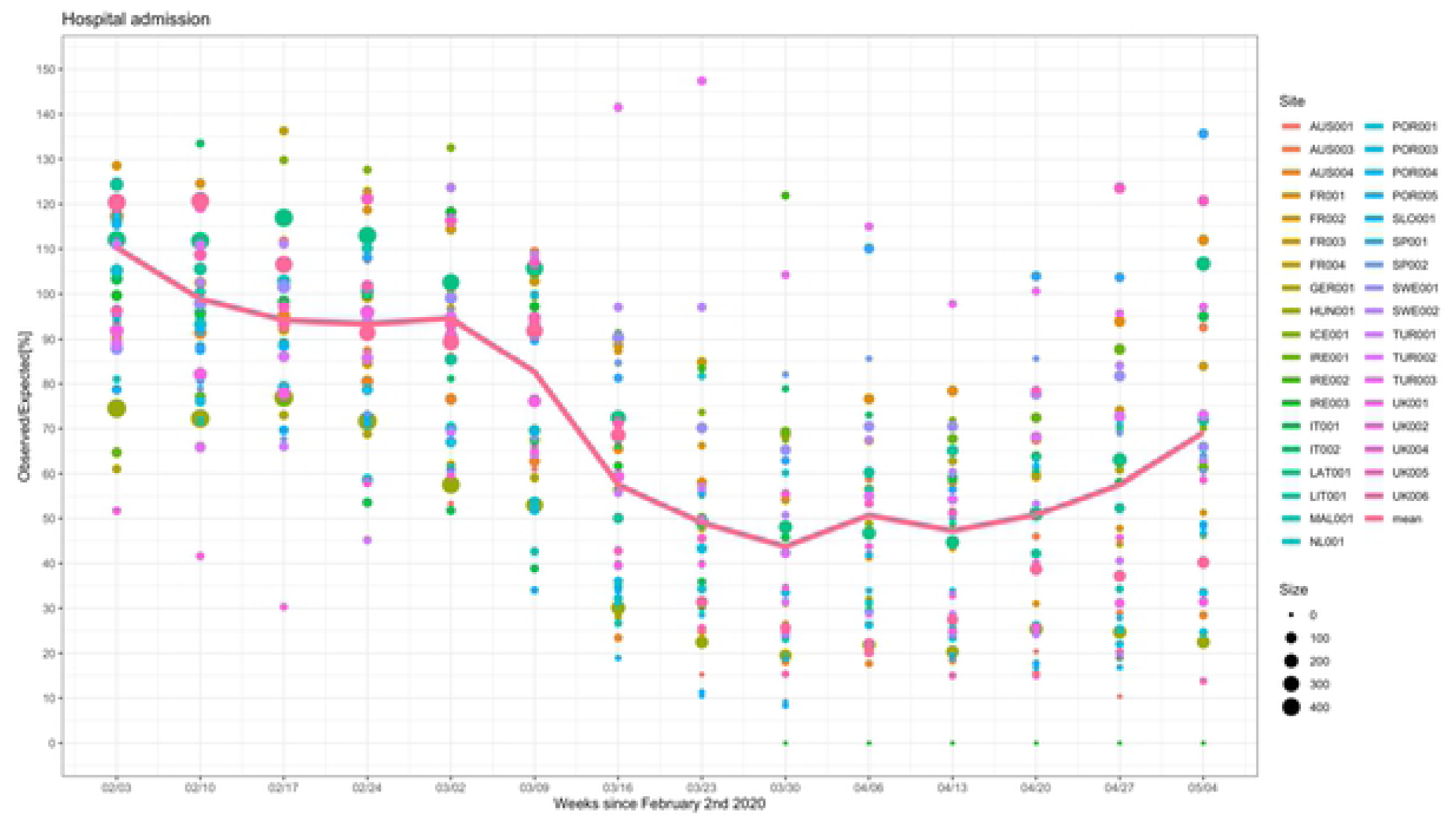
Observed and predicted in % for the number of hospital admissions for patients attending the emergency department. The observed and predicted number of children admitted to hospital from the emergency department in countries across Europe in the weeks following February 2^nd^ 2020 until May 11^th^ 2020, for all sites combined. The colour and the size of the dots reflect the actual number of ED attendances for each site and for each time window. The line connects the mean of the observed vs predicted point estimates for each of the individual sites for each time window.

**Fig 4.**
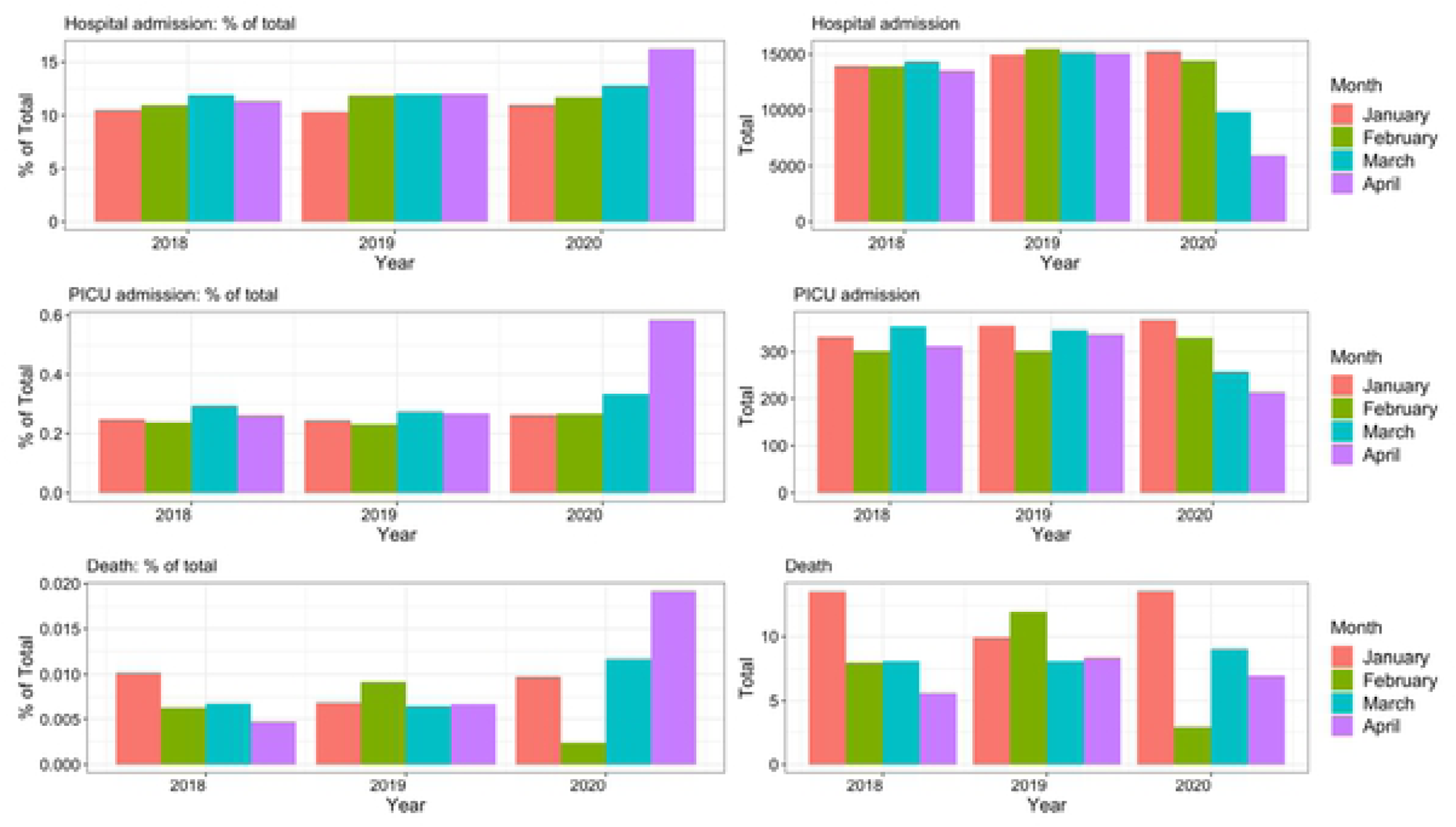
The 28-day mean number of hospital admissions, intensive care admissions and deaths in the emergency department for the period January – April over a three-year period. Percentages of total ED attendances (left) and absolute numbers (right) of children admitted to hospital (top), pediatric intensive care units (middle), or died in the ED (bottom); comparing the 28-day mean numbers for the months of January – April for 2018 vs. 2019 vs. 2020.

### Diagnoses

The 28-day mean numbers for common communicable diseases decreased in absolute and relative frequencies (Table 3, Fig 5), in particular for tonsillitis, otitis media, gastro-intestinal infectionsand LRTIs. Decreases were also seen in common childhood injuries such as minor head injuries and radius fractures. No increase in absolute numbers were seen for several uncommon diagnoses suggested to be linked with SARS-CoV-2 infection, such as diabetic ketoacidosis, intussusceptionand testicular torsion, even when stratified for high SARS-CoV-2 prevalence countries (S16 Fig). Mental health attendances declined during the first phase of the COVID-19 pandemic in absolute terms, but this corresponded with an increase in relative frequency. Fig 6, reflecting the observed vs predicted numbers for the eight selected diagnoses, shows that the impact on appendicitis was lower than on the other diagnoses groups. Mental health issues, radius fracturesand minor head injuries were all affected, but there was evidence that attendances increased from the end of March. In contrast, attendances for LRTI, otitis media, gastro-intestinal infections and tonsilitis remained low. Poisson models showed no significant difference between mental health, minor head trauma and radius fracture. There was evidence of significant difference between infections and trauma and mental health, with bigger reductions in infections. When communicable diseases were combined, there was a clear difference between surgical presentation (appendicitis), communicable diseases and ‘other’ (mental health, radius fracture and head trauma) (Table 2).

**Fig 5.**
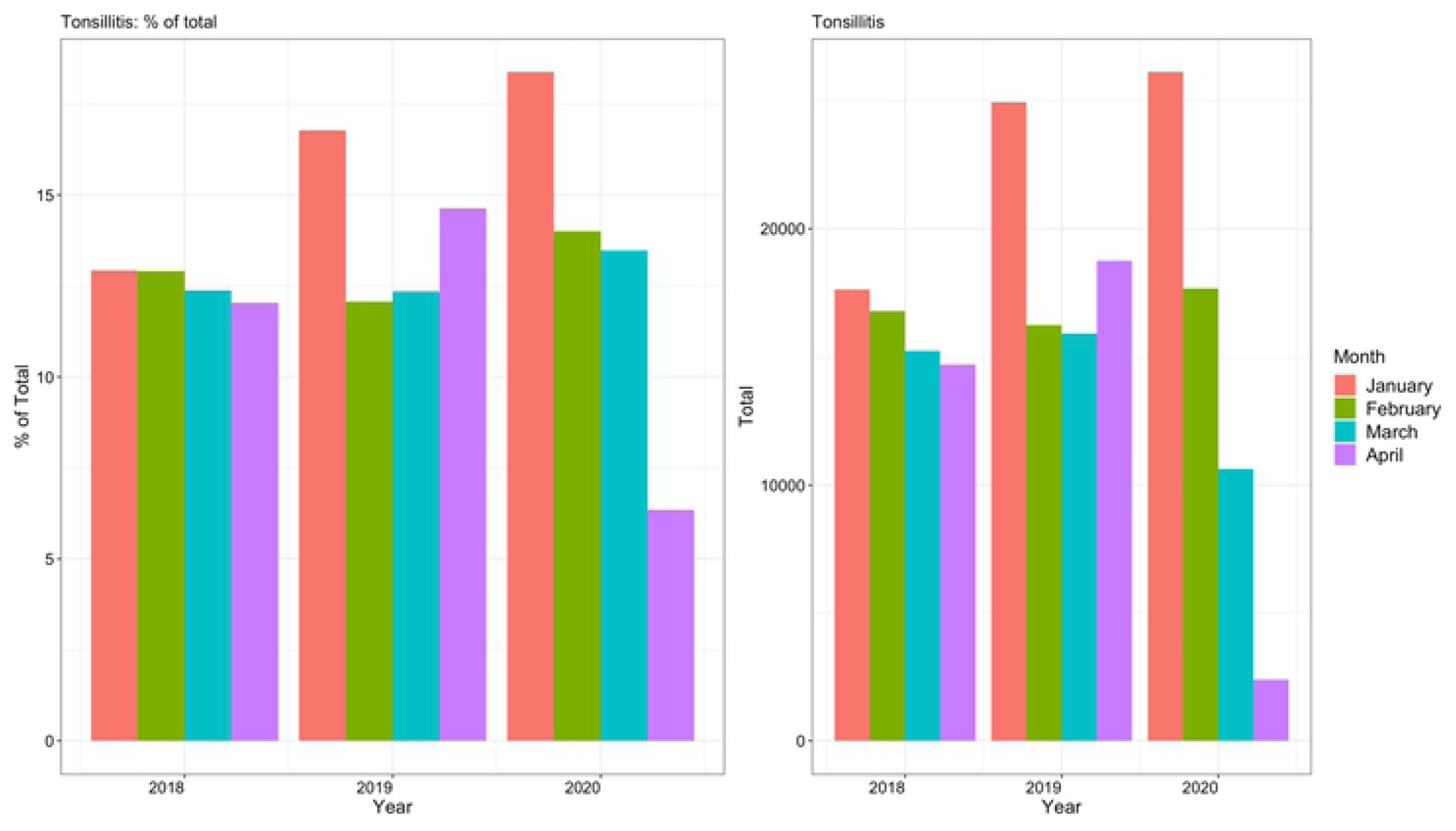
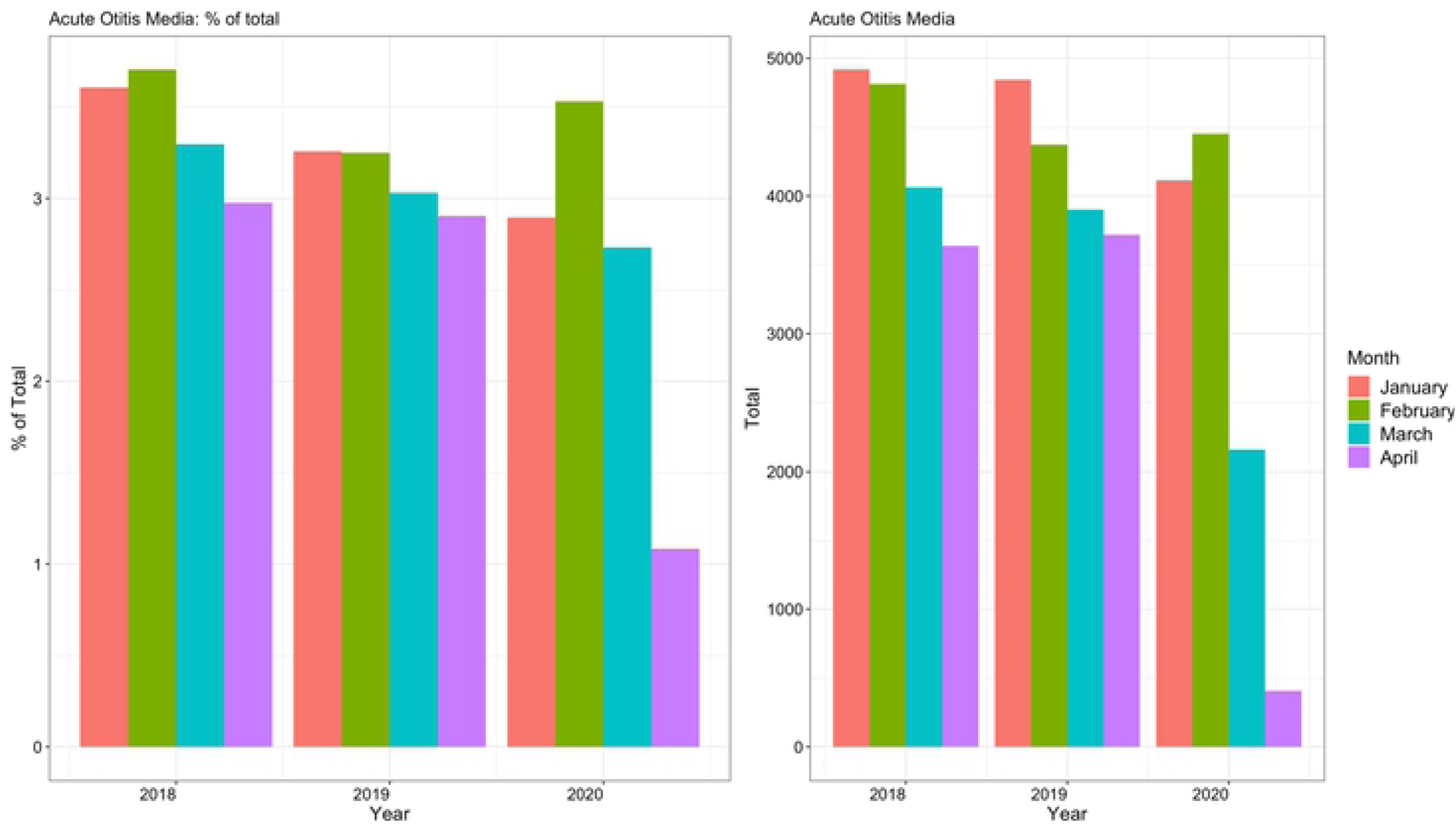
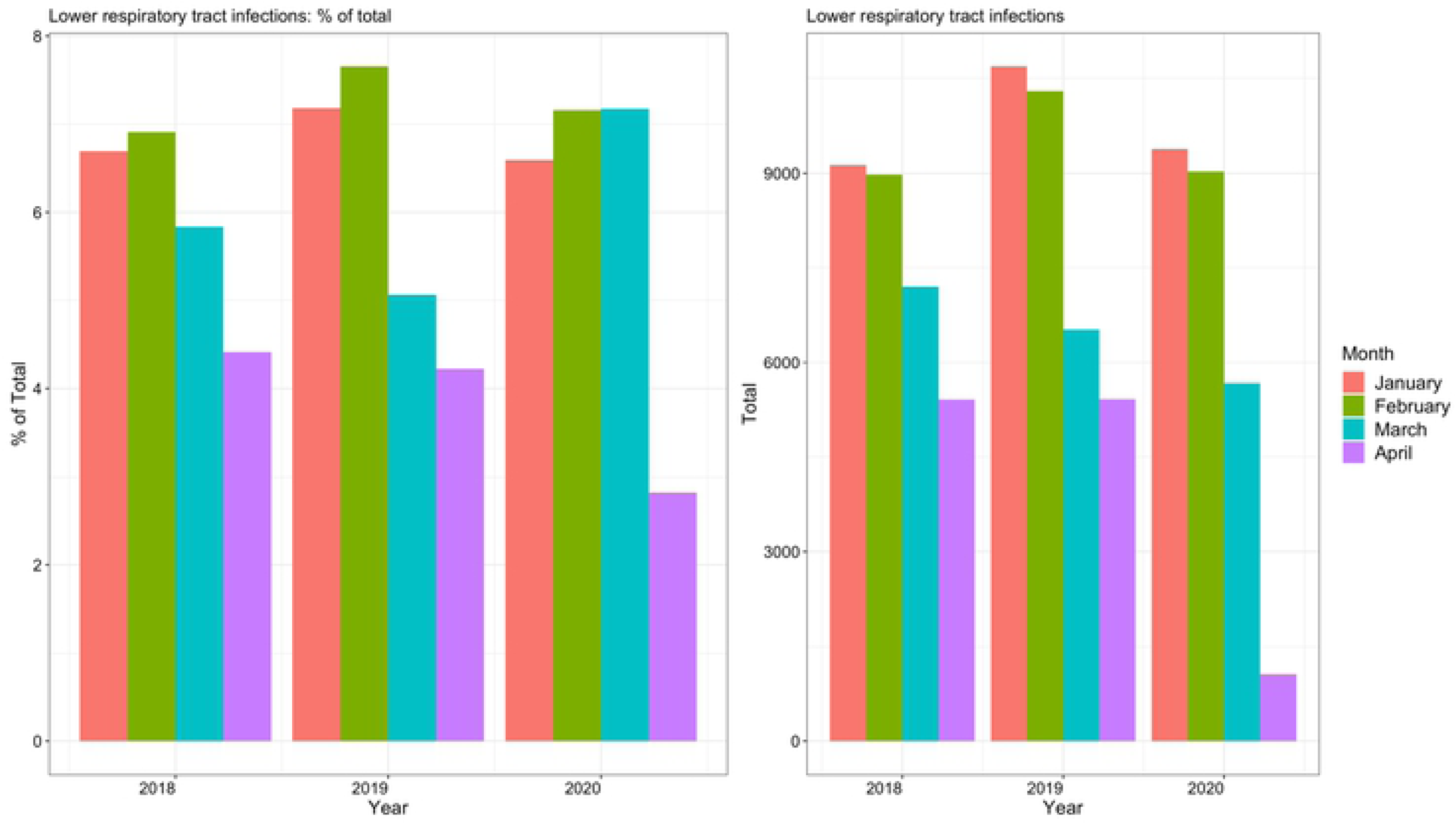
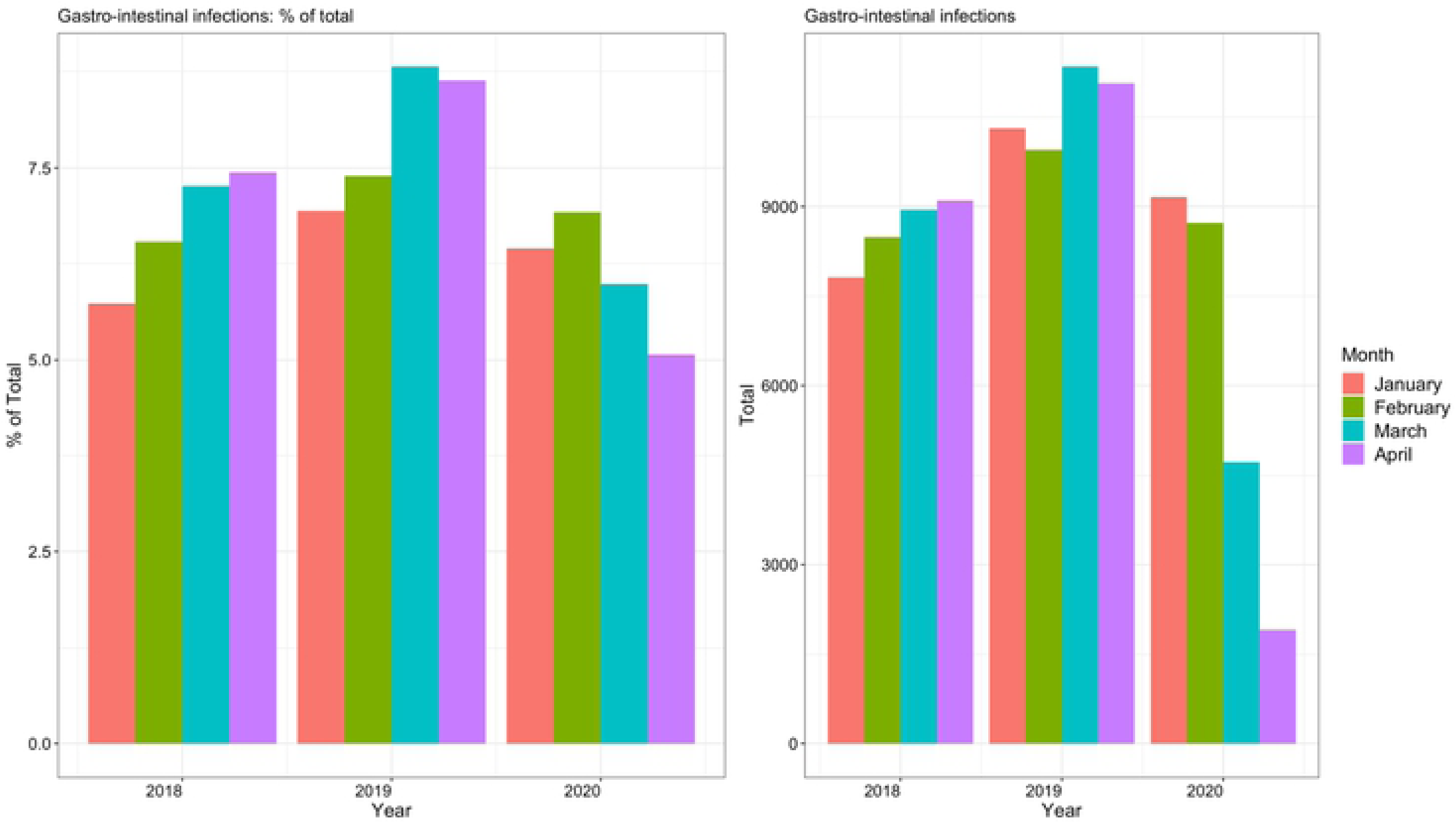
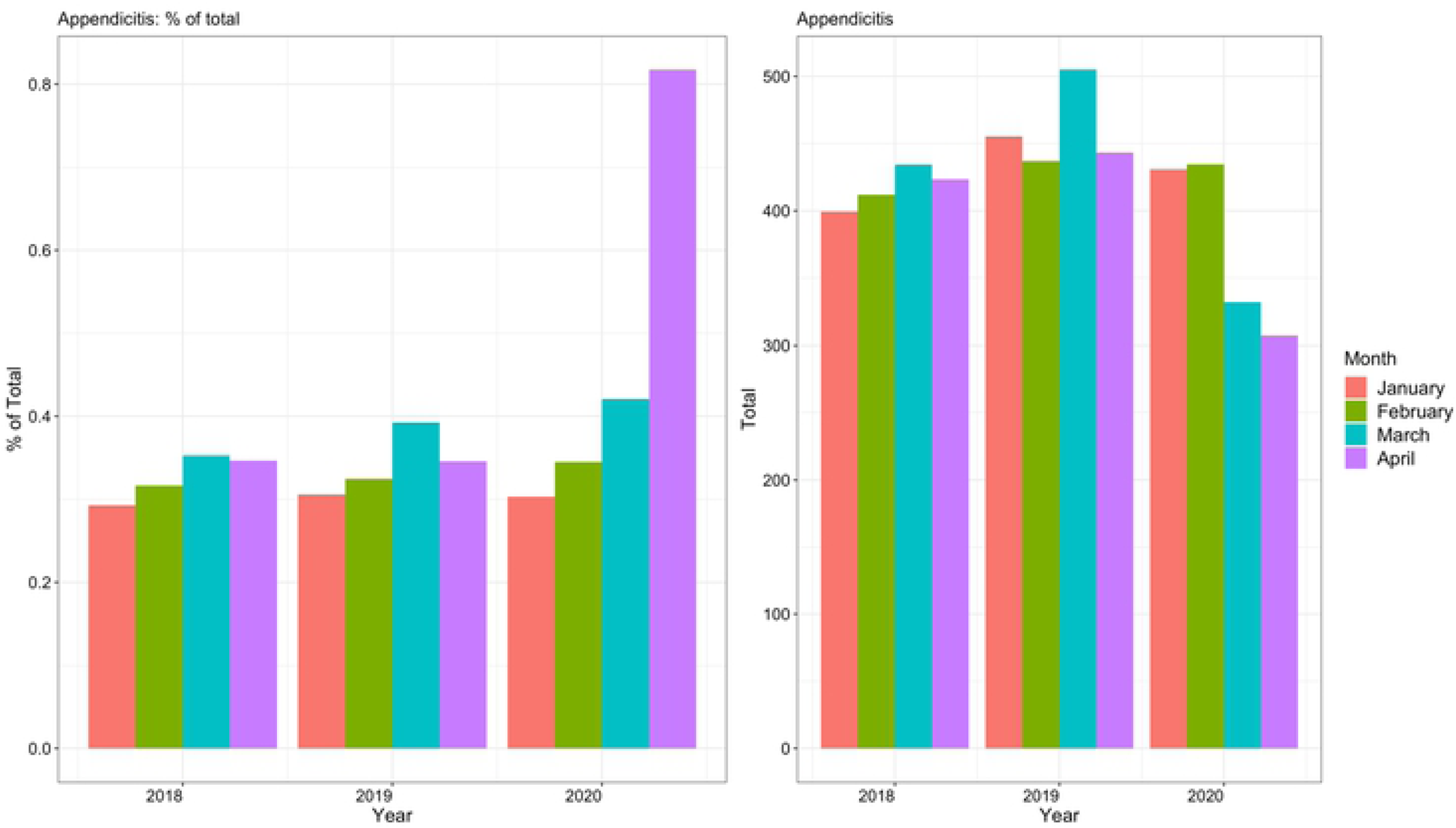
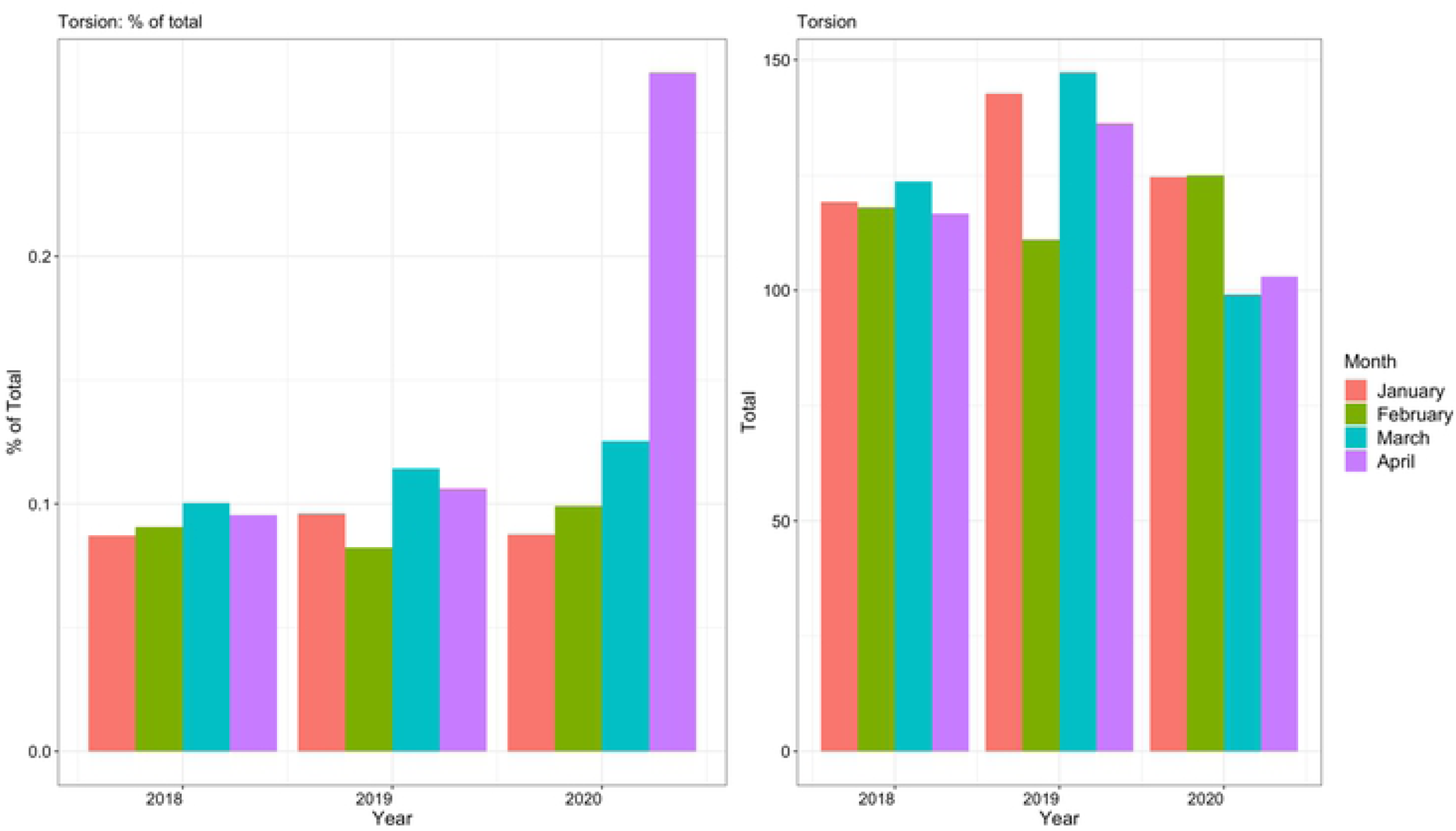
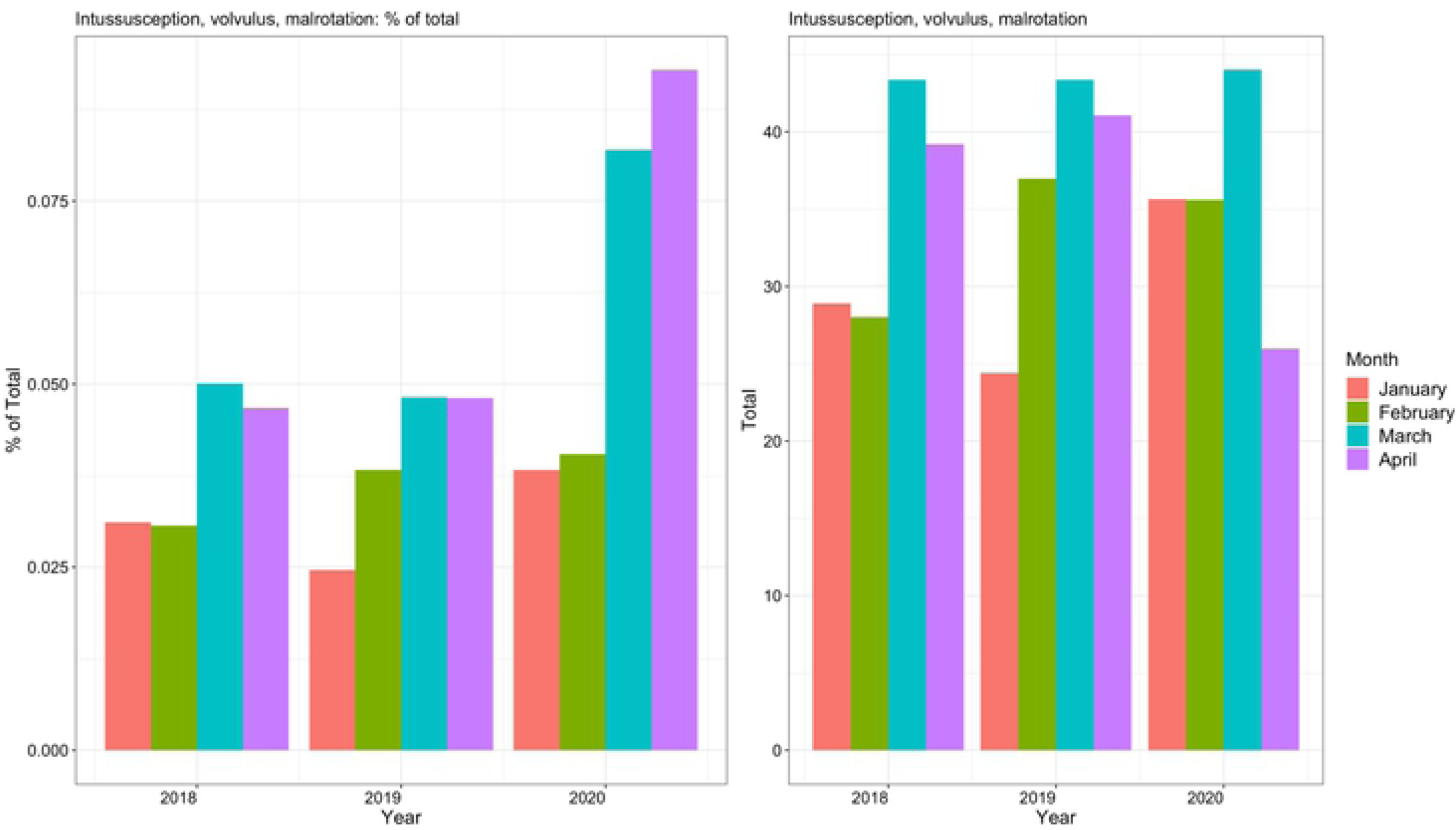
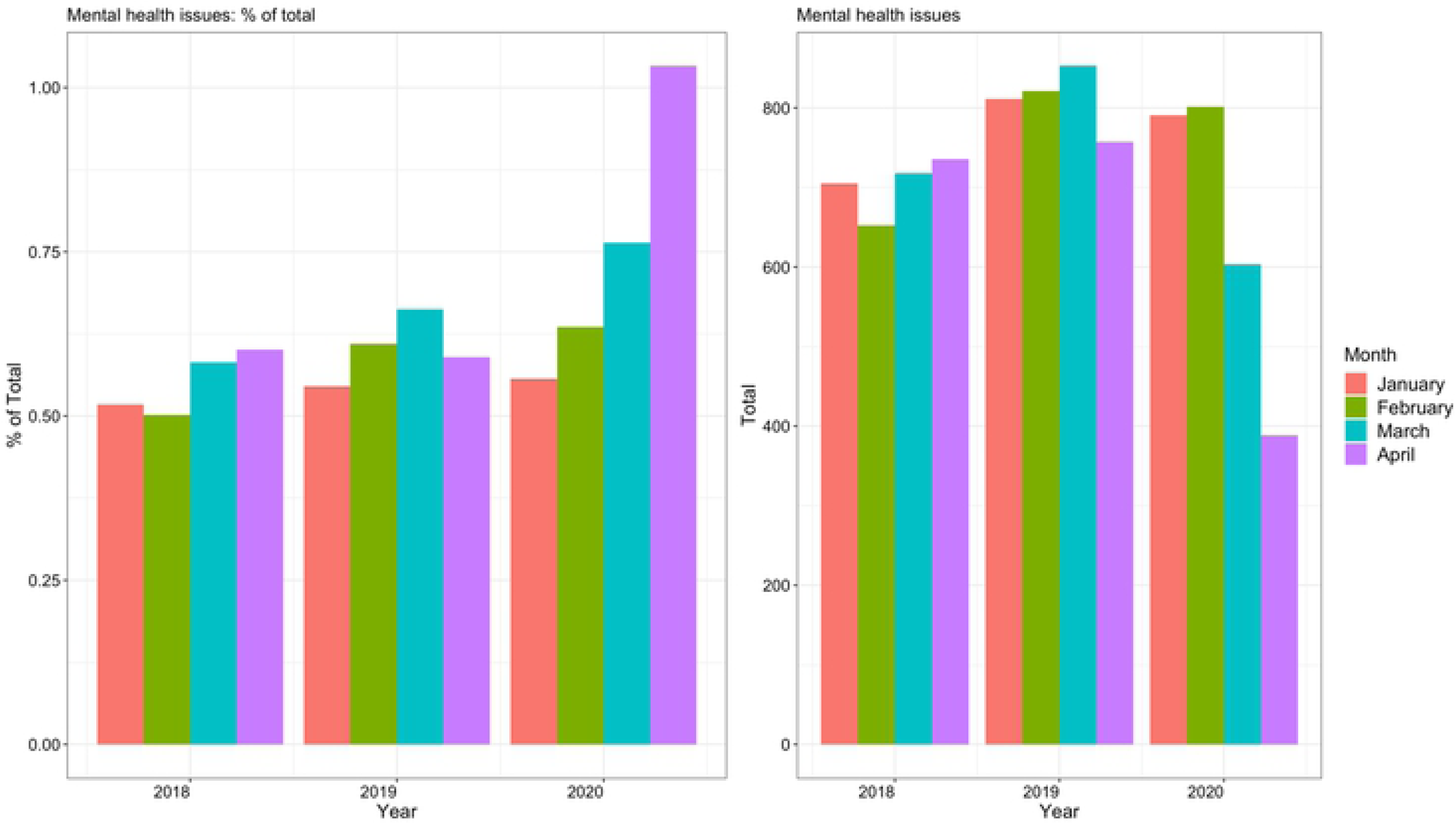
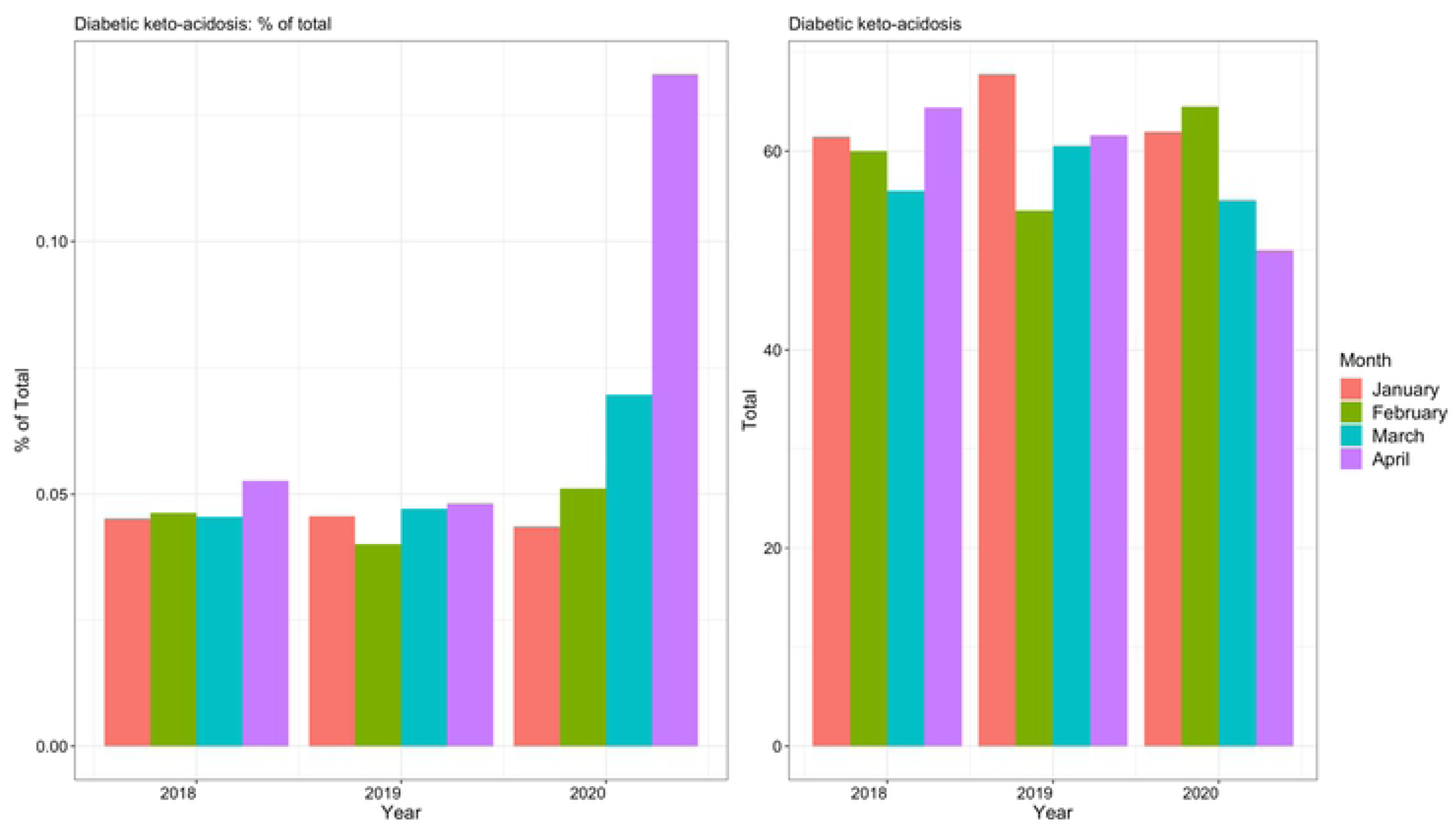
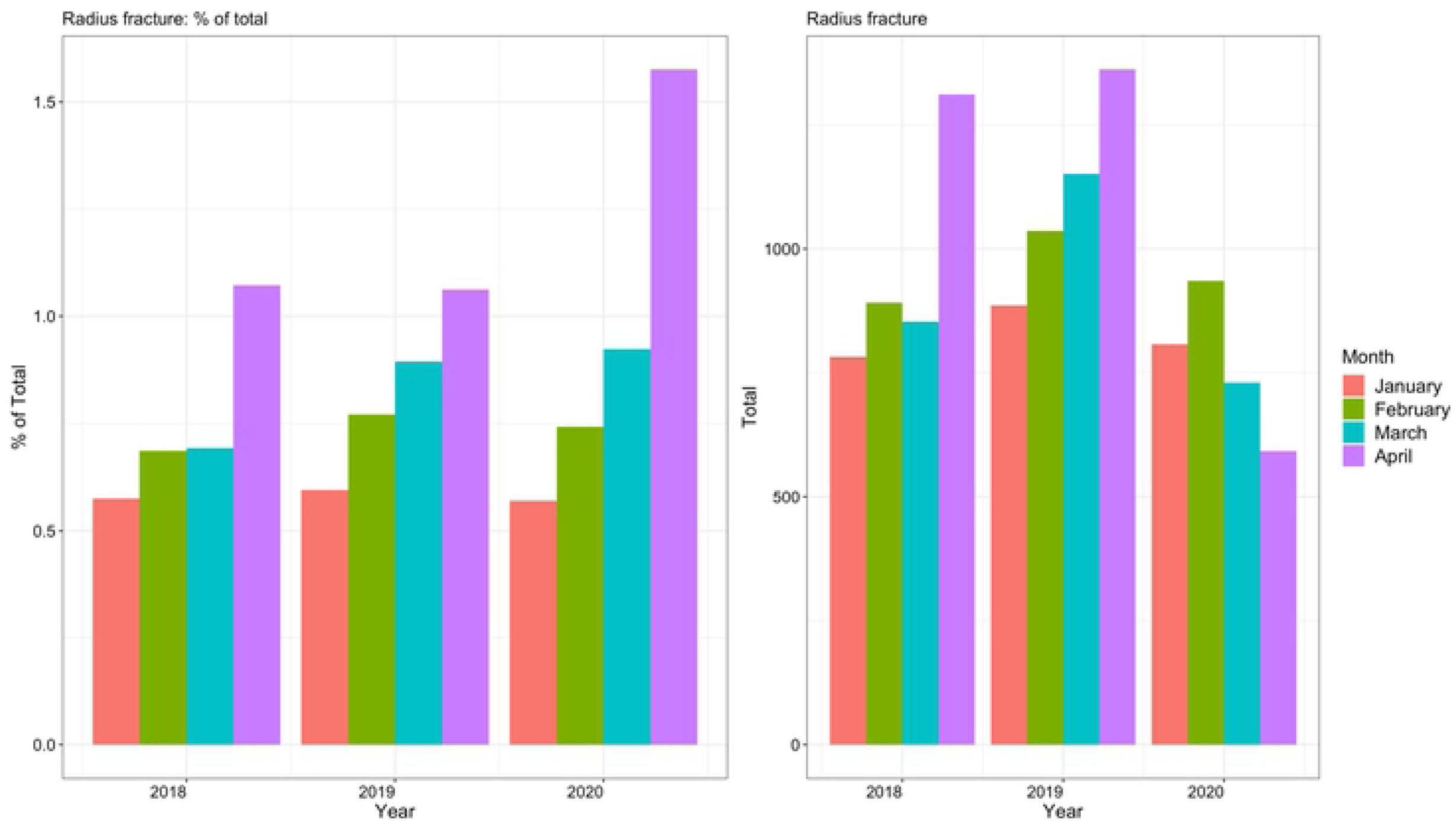
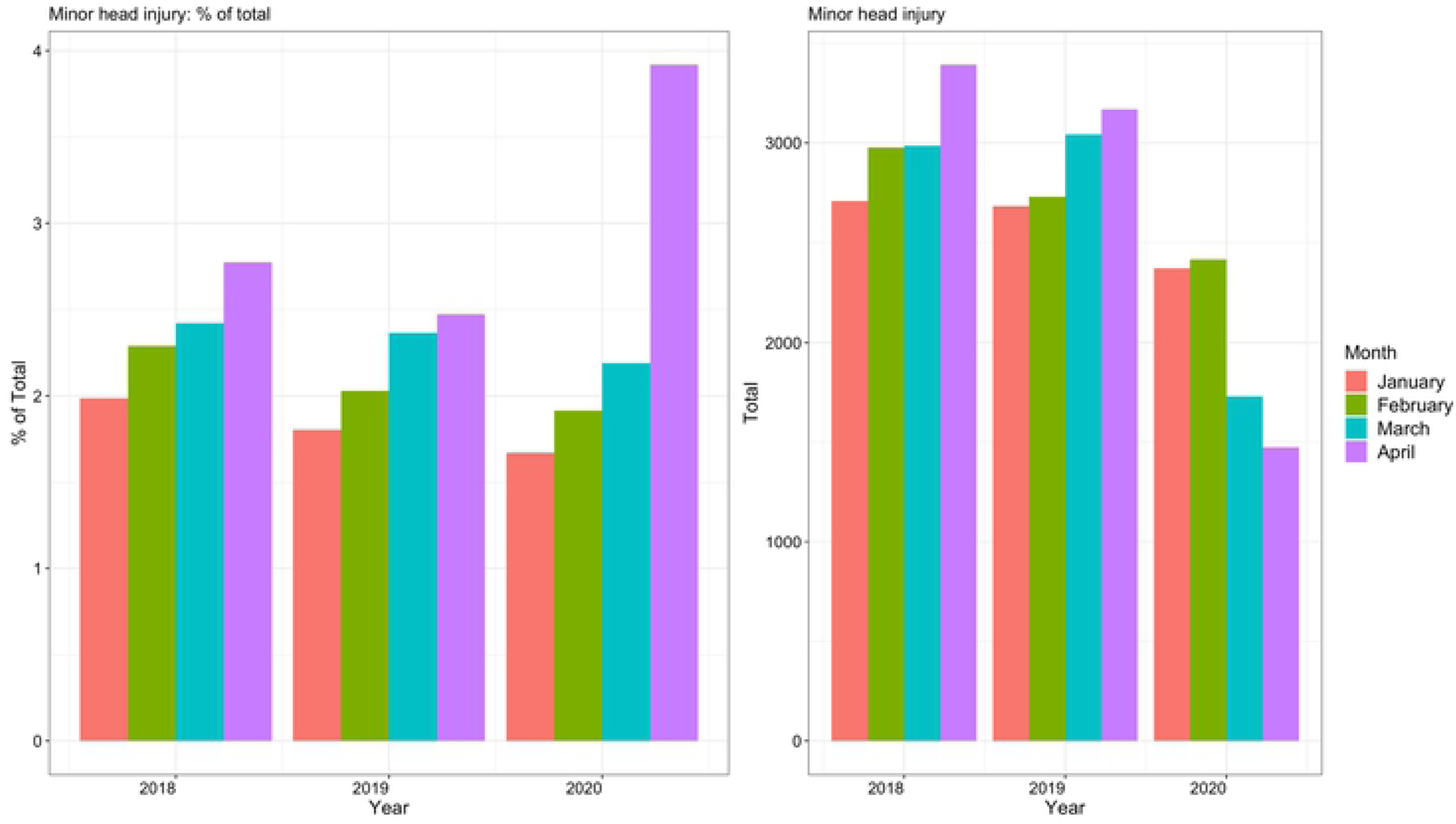
The 28-day mean number of selected clinical diagnoses in the emergency department for the period January – April over a three-year period. Percentages of total ED attendances (left) and absolute numbers (right) of children with diagnosis of a) tonsillitis, b) otitis media, c) lower respiratory tract infections (LRTI), d) gastro-intestinal (GI) infections, e) appendicitis, f) testicular torsion, g) intussusception, volvulus and malrotation (combined group), h) mental health issues, i) diabetic keto-acidosis, j) radius fracture, k) minor head injury; comparing the 28-day mean numbers for the months of January – April for 2018 vs. 2019 vs. 2020.

**Fig 6.**
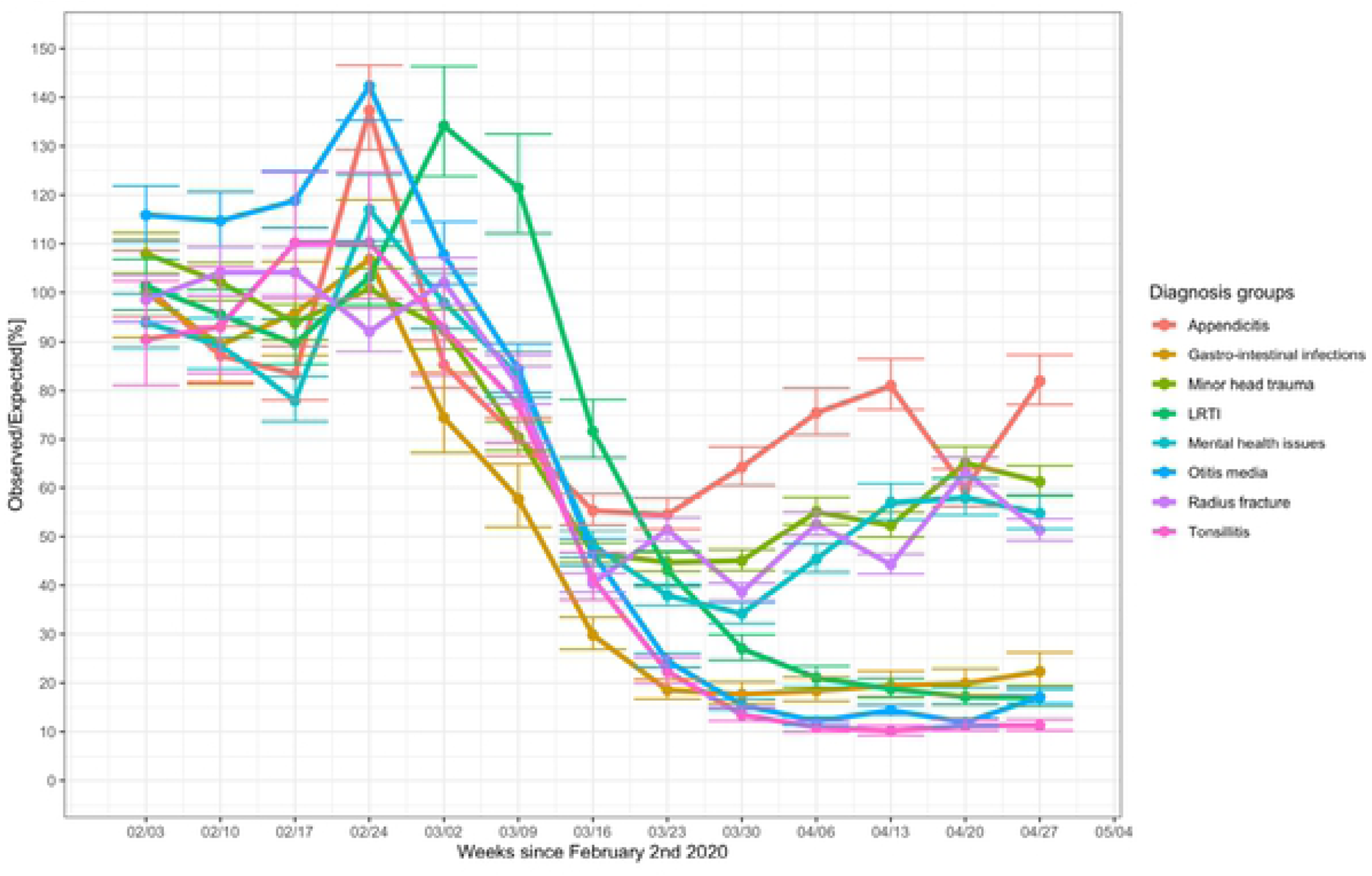
Observed and predicted in % for the number of selected diagnoses. The observed and predicted numbers of eight selected diagnoses for all sites combined, for the period following February 2^nd^ 2020 until May 4^th^ 2020.

**Table 3.** The 28-day mean numbers of selected clinical diagnoses in the emergency department for the period January – April over a three-year period. Absolute numbers (and % of total children seen in the ED) of children with diagnosis of a) tonsillitis, b) otitis media, c) lower respiratory tract infections, d) gastro-intestinal infections, e) appendicitis, f) testicular torsion, g) intussusception, volvulusand malrotation, h) mental health issues, i) diabetic keto-acidosis, j) radius fracture, k) minor head injury; comparing the 28-day mean numbers for the months of January – April for 2018 vs. 2019 vs. 2020.

|  | 2018 |  |  |  | 2019 |  |  |  | 2020 |  |  |  |
| --- | --- | --- | --- | --- | --- | --- | --- | --- | --- | --- | --- | --- |
|  | January | February | March | April | January | February | March | April | January | February | March | April |
| Tonsillitis | 17627<br>(12·9%) | 16781<br>(12·9%) | 15249<br>(12·4%) | 14722<br>(12·0%) | 24939<br>(16·8%) | 16231<br>(12·1%) | 15898<br>(12·3%) | 18751<br>(14·6%) | 26118<br>(18·4%) | 17668<br>(14·0%) | 10639<br>(13·5%) | 2382<br>(6·3%) |
| Otitis Media | 4917<br>(3·6%) | 4814<br>(3·7%) | 4063<br>(3·3%) | 3639<br>(3·0%) | 4843<br>(3·3%) | 4370<br>(3·2%) | 3900<br>(3·0%) | 3719<br>(2·9%) | 4115<br>(2·9%) | 4452<br>(3·5%) | 2156<br>(2·7%) | 407<br>(1·1%) |
| Lower respiratory tract infections | 9129<br>(6·7%) | 8981<br>(6·9%) | 7197<br>(5·8%) | 5407<br>(4·4%) | 10684<br>(7·2%) | 10294<br>(7·7%) | 6519<br>(5·1%) | 5415<br>(4·2%) | 9374<br>(6·6%) | 9028<br>(7·2%) | 5667<br>(7·2%) | 1059<br>(2·8%) |
| Gastro-intestinal infections | 7809<br>(5·7%) | 8488<br>(6·5%) | 8946<br>(7·3%) | 9103<br>(7·4%) | 10313<br>(6·9%) | 9949<br>(7·4%) | 11345<br>(8·8%) | 11066<br>(8·6%) | 9158<br>(6·4%) | 8726<br>(6·9%) | 4719<br>(6·0%) | 1905<br>(5·1%) |
| Appendicitis | 399<br>(0·3%) | 412<br>(0·3%) | 434<br>(0·4%) | 424<br>(0·3%) | 455<br>(0·3%) | 437<br>(0·3%) | 505<br>(0·4%) | 443<br>(0·3%) | 431<br>(0·3%) | 435<br>(0·3%) | 332<br>(0·4%) | 307<br>(0·8%) |
| Testicular torsion | 119<br>(0·1%) | 118<br>(0·1%) | 124<br>(0·1%) | 117<br>(0·1%) | 143<br>(0·1%) | 111<br>(0·1%) | 147<br>(0·1%) | 136<br>(0·1%) | 125<br>(0·1%) | 125<br>(0·1%) | 99<br>(0·1%) | 103<br>(0·3%) |
| Intussusception, volvulus and malrotation | 33<br>(0·0%) | 37<br>(0·0%) | 56<br>(0·0%) | 58<br>(0·0%) | 40<br>(0·0%) | 58<br>(0·0%) | 63<br>(0·0%) | 60<br>(0·0%) | 55<br>(0·0%) | 44<br>(0·0%) | 53<br>(0·1%) | 30<br>(0·1%) |
| Mental health conditions | 705<br>(0·5%) | 652<br>(0·5%) | 718<br>(0·6%) | 736<br>(0·6%) | 811<br>(0·5%) | 821<br>(0·6%) | 853<br>(0·7%) | 757<br>(0·6%) | 791<br>(0·6%) | 802<br>(0·6%) | 603<br>(0·8%) | 388<br>(1·0%) |
| Diabetic keto-acidosis | 61<br>(0·0%) | 60<br>(0·0%) | 56<br>(0·0%) | 64<br>(0·1%) | 68<br>(0·0%) | 54<br>(0·0%) | 61<br>(0·0%) | 62<br>(0·0%) | 62<br>(0·0%) | 64<br>(0·1%) | 55<br>(0·1%) | 50<br>(0·1%) |
| Radius fracture | 783<br>(0·6%) | 891<br>(0·7%) | 853<br>(0·7%) | 1311<br>(1·1%) | 886<br>(0·6%) | 1036<br>(0·8%) | 1153<br>(0·9%) | 1362<br>(1·1%) | 809<br>(0·6%) | 935<br>(0·7%) | 730<br>(0·9%) | 592<br>(1·6%) |
| Minor head injury | 2709<br>(2·0%) | 2976<br>(2·3%) | 2985<br>(2·4%) | 3392<br>(2·8%) | 2682<br>(1·8%) | 2730<br>(2·0%) | 3043<br>(2·4%) | 3169<br>(2·5%) | 2372<br>(1·7%) | 2415<br>(1·9%) | 1728<br>(2·2%) | 1472<br>(3·9%) |

### Sensitivity analyses

The sensitivity analyses for the Poisson modelling without TUR003 resulted in IRRs slightly nearer to one, meaning all associations were slightly weaker. The impact on the coefficient for tonsillitis was notable, increasing the IRR from 0.19 (95% CI 0.17 to 0.21) to 0.37 (95% CI 0.34 – 0.41). There was also a notable impact on the difference between PICU admissions and admissions in general, which were less significant when TUR003 was removed.

## Discussion

Reductions in the numbers of children attending EDs were consistently seen across Europe during the first phase of the COVID-19 pandemic. There was variation between countries, but within countries patterns were similar. The levels to which ED attendances decreased appeared to be related to the introduction of infection prevention measures, changes made to local health systemsand national SARS-CoV-2 prevalence. Reduced ED attendances were seen for all age groups, with smaller reductions in children aged below 1 year. The impact was largest and sustained for communicable diseases, whereas other groups of diagnoses trended towards normal levels of ED attendances by the end of the study period after initial reduced ED attendance rates.

Our findings of reduced pediatric ED attendances are consistent with other studies from around the world. [6–10,22–24] The observed reduction in ED attendances will likely be multifactorial. For example, children with asthma often frequent EDs, but they had fewer exacerbations needing ED visits during the first phase of the COVID-19 pandemic. Proposed reasons include reduced air pollution, reduced social mixing with exposures to viral trigger, and improved compliance with medication at home. [25,26]

Earlier studies suggested that infection prevention measures may have resulted in delayed presentations to hospitals. [11,12][27–29] In our study, children with more severe conditions, as measured by triage urgency, need for hospital admission and PICUand death, continued to attend hospital more frequently compared to those with minor injuries and illnesses, although overall absolute numbers fell. This was in line with other studies reporting similar reductions in children with high triage urgency or need for hospital admission. [6,30–38]

Defining the harm of delayed presentations, as well as establishing what contributed to a possible delay, can be difficult. [39] In an attempt to distinguish the delay in seeking care from harm sustained, Roland et al. concluded that only a minority (6 out of 51 (11.8%)) of children with a potential delay in presentation were admitted to one of seven hospitals. [40] Contradictory conclusions have been reported for the delay in presentations and for potential harm sustained for diagnoses of appendicitis [41,42] and testicular torsion [43–46], portraying a picture that organising regional health care delivery is important to ensure continued access to pediatric urgent and emergency care during a pandemic. In addition, our data suggest that, despite overall falling ED attendances, presentations requiring surgical interventions remained stable, reiterating that access to surgical teams and the ability to perform emergent surgical procedures are crucial.

Evidence is mounting that SARS-CoV-2 is directly involved in the pathogenesis of new onset diabetes. [47,48] Unsworth et al. first reported an increase of new onset type 1 diabetes in children and a possible link with SARS-CoV-2 in the UK. [15] Additional cohort studies found divergent associations between SARS-CoV-2, new onset diabetesand decompensation of pre-existing diabetes. [49–52] Our data did not identify increased incidence of diabetic keto-acidosis during the first phase of the COVID-19 pandemic. It might well be that clusters of new onset diabetes can be found in high prevalence areasand that we failed to capture this in our study. Likewise, if SARS-CoV-2 acts as a precipitator, there might be a delay in the manifestation of new onset diabetes, and with reduced prevalence of typical viral triggers, this increase might only become apparent later in the pandemic. [53] We were not able to differentiate between new onset diabetes and decompensation of pre-existing diabetes.

We found a reduction of children with mental health conditions presenting to the EDs during the first phase of the COVID-19 pandemic in Europe, similar to findings from studies elsewhere. [8,54–57] This is unlikely to reflect the considerable mental health issues encountered in the wider pediatric and adolescent populations [58] and of the experiences later in the pandemic, with, amongst others, reported increases in eating disorders in children and young people. [59] Joyce et al observed an overall decrease in mental health issues in ED, albeit an increase in self harm and deliberate ingestions. [60] Despite the reduction in absolute numbers, there was an increase in the proportion of attendances attributable to mental health potentially contributing to the heightened awareness for mental health issues in the first COVID-19 wave.

Prior to the current COVID-19 pandemic, limited data were available describing the impact of infection prevention measures on urgent and emergency pediatric care in high income countries. One study found a decrease in respiratory infections of 42%and decreased ED attendances of 28% following school closures for an influenza outbreak in Israel. [61] Similar patterns were seen during the SARS outbreak in 2003, [62–64] and the MERS outbreak in 2015. [65] These studies also reported a larger impact in reduced ED utilisation for children than for adults. In contrast, the H1N1 influenza pandemic in 2009 generally led to increased ED utilisation, with higher levels of acuity. [66–68] One previous study had reported reduced pediatric ED attendance rates for flu like illness and respiratory tract infections following school closures. [69] Another study reported increased pediatric ED attendance numbers following media reports on health threats of the H1N1 virus. [70] Altogether, previous evidence of infectious disease outbreaks suggests a similar impact on pediatric urgent and emergency care following the introduction of public health and infection prevention measures. However, this is to a lesser extent than what was observed with the COVID-19 pandemic, and one that is dependent on childhood susceptibility for the infectious pathogen.

### Strengths and limitations

Our study presents multinational data enabling the comparison between infection prevention measures, national SARS-CoV-2 prevalenceand the impact on acute illness and injuries in children between European countries. Most participating sites were tertiary institutions, with dedicated pediatric emergency medicine teams, with potential implications for the generalisability of our findings. At present, no standardized data extraction system for pediatric urgent and emergency care exists between European countries; and the EPISODES study is the first to navigate the difficulties of dealing with different data systems, data availabilityand varying coding practices. Hence, also limited by the time restrictions caused by the COVID-19 pandemic, some sites were not able to provide data for all domains, and two sites (NL002, HUN002) were only able to provide data for part of the study duration.

Limitations of electronic health records to describe patients’ diagnoses are well known. [71] Participating study sites had unique coding systems, and we urged all study teams to be consistent in transforming local data to fit the study clinical report form.

Although most diagnoses linked to SARS-CoV-2 in children were included in the pre-defined clinical report form, other diagnoses might be of interest in future studies. Of note, coding for children with Multi Inflammatory Syndrome in Children (MIS-C) proved unreliable, with no unique diagnostic codes available for this new disease in automated coding systems.

As the data were collected in aggregated form, thereby negating some of the difficulties with data protection regulations, we were not able to stratify for severity of specific diagnosis or age groups. We observed large differences between sites for the number of annual ED attendances and the number of patients with high triage urgency and hospital admissions, reflecting both case-mix and diversity of patient management. We analysed data mostly on a site-by-site basis, by using predicted versus observed ratios, and thus dealing with heterogeneity between sites.

We used the cumulative 14-day rate of new cases per 100,000 of the population to identify high prevalence countries, but indications for SARS-CoV-2 testing differed between countries, and this could have led to under- or overestimation of national prevalence rates. Moreover, national prevalence numbers might wrongly reflect any regional variation, but, for example, in the UK, identical patterns in ED attendances were seen across the five sites, despite large variations in SARS-CoV-2 prevalence during the first phase of the COVID-19 pandemic (S17 Fig).

## Conclusion

Reductions in overall ED attendances were seen across our study sites during the first phase of the COVID-19 pandemic, with health systems across Europe impacted similarly. In most sites, there was no suggestion of disproportionate numbers of more severely unwell children. In the first phase of the COVID-19 pandemic, the relative increase in cases of diabetic keto-acidosis or mental health issues might have contributed to a biased perception about increased occurrence, yet this is not supported by an increase in absolute numbers of cases in our data. Our study informs how pediatric emergency medicine can prepare for future pandemics, taking into account that different infectious diseases outbreaks can affect children differently, and illustrates the potential of electronic health records to monitor trends in urgent and emergency care for children.

## Data sharing statement

All data used for this manuscript will be made available to others immediately following the time of publication with no end date.

## Author contributions

The principal investigator for the EPISODES study is RN. RN, LT conceived and planned the study and worked alongside the EPISODES Steering Committee to establish the protocol and data collection for this study. RN, IM obtained sponsorship and ethical consent. RN, KH, SB, RO, HM, DB, ZB, RF, DR, HM were responsible for coordinating the study and approaching and training study sites; RO, SB, RN led on gaining REPEM network support. All authors were responsible for local data access, data collection, data cleaning and modificationand obtaining the local approvals. RN, KH, SB, RO, HM, DB, ZB, RF, DR, HM were responsible for data analysis and interpretation. RN was responsible for writing the first draft of this manuscript with RF, KR, KH forming the initial writing group. All authors, as part of the EPISODE Steering group provided critical feedback and helped shape the research, analysis and manuscript. All authors had full access to the data, and all authors commented on and approved of the final version of this manuscript as submitted. RGN and KH verified all underlying data and act as guarantors for this study.

## Data Availability

All study data are available from via URL XXX

## Acknowlegdements

Celia Nekrouf (Pediatric Emergency Department, Hopital Universitaire Robert-Debre, Paris, France); Marcello Covino (Emergency Department, Fondazione Policlinico Universitario A. Gemelli IRCCS, Rome, Italy); Benita Lund (Medical Secretary; Pediatric emergency department, Sachs’ Children and Youth Hospital, Stockholm, Sweden); Izabella Lottiger (Pediatric Emergency department, Astrid Lindgren Children Hospital, Stockholm, Sweden); Sharna Crosdale (Contracts office; Imperial College London, UK); Sarah Sheedy (Research nurse; Emergency Department, Bristol Royal Hospital for Children, Bristol, UK); James Allbones (Information & Performance Analyst; Birmingham Women’s and Children’s NHS Foundation Trust, UK), William Jones (University Hospitals of Leicester NHS Trust, UK); Frazer Snowdon and Matthew Ryan (Pediatric emergency department, Alder Hey Children’s NHS Foundation Trust, Liverpool, UK), Carlos Saiz-Hernando (IT analyst; Department of Medical Documentation, Cruces University Hospital, Bilbao, Spain); Ellen Barry (research nurse; National Children’s Research Centre, Dublin, Ireland) and Fiona Leonard (data analyst, Children’s Health Ireland, Ireland); Ernst Eigenbauer (data analyst) and Katharina Lieb (medical student; Department of Pediatrics and Adolescent Medicine, Vienna, Austria); Sanne Vrijland (medical student; Erasmus MC – Sophia Children’s hospital, Rotterdam, The Netherlands); Catarina Cordeiro (Pediatric Emergency Service, Hospital Pediátrico, Centro Hospitalar e Universitário de Coimbra, Portugal); Mark Camenzuli (Senior systems administrator; Mater Dei Hospital, Malta); Sandra Distefano (Clinical Performance Unit, Mater Dei Hospital, Malta); Karin Kittl-Mitteregger (HIS Management and Clinical Processes, Paracelsus Medical University, Salzburg, Austria); Marta Arpone (Research Assistant, University of Padova, Italy)

## Supporting information

**S1 Table. Overview of participating study sites**

**S2 File. Clinical report form**

**S3 Table. List of time windows for data entry**

**S4 Fig. Spiderplot for availability of data**

**S5 Table. ICD-10 Guidance for coding of diagnosis**

**S6 Table. List of national social distancing measures**

**S7 Table. List of national SARS-CoV-2 rates**

Legend:

The dates and numbers of SARS-CoV02 infections in each of the study sites’ countries participating in. the EPISODES study.

**S8 Table. List of approvals**

**S9 Fig. Annual emergency department attendance numbers in 2018 and 2019**

Legend:

The total number of pediatric emergency departments attendances for each of the study sites for 2018 and 2019 (pre-COVID-19). Overall, the numbers of ED attendances in 2018 and 2019 were similar for each of the study sites, with considerable diversity between the study sites and TUR003 seeing more children in their ED than the other study sites.

**S10 Fig. Observed and predicted in % for the number of children for different age categories attending the emergency department**

Legend:

The observed and predicted number of children presenting to emergency departments in countries across Europe in the weeks following February 2^nd^ 2020 until May 11^th^ 2020, for all sites combined, for children a) aged 0-1 years, b) 1-2 years, c) 2-5 years, d) 5-12 years, e) 12 – 16 years. The colour and the size of the dots reflect the actual number of ED attendances for each site and for each time window. The line connects the mean of the observed vs predicted point estimates for each of the individual sites for each time window.

**S11 Fig. Observed and predicted in % for the number of children for different age categories attending the emergency department for individual sites**

**S12 Fig. The observed and predicted (in %) of total ED attendances for each country.**

Legend:

The observed and predicted number of children presenting to emergency departments in countries across Europe for which data from only one study site were available in the weeks following February 2^nd^ 2020 until May 11^th^ 2020. A timeline is plotted (dashed line) to show the dates of the introduction of national social distancing measures.^1^ One site from Netherlands and one site from Hungary were excluded from these analyses as these sites could not provide data for the entire study duration.

**S13 Fig. Observed and predicted in % for different triage categories of children attending the emergency department**

Legend:

The observed and predicted number of children presenting to emergency departments in countries across Europe in the weeks following February 2^nd^ 2020 until May 11^th^ 2020, for all sites combined, for children a) non urgent and standard triage classification, b) urgent triage classification, c) emergency and very urgent triage classification. The colour and the size of the dots reflect the actual number of ED attendances for each site and for each time window. The line connects the mean of the observed vs predicted point estimates for each of the individual sites for each time window. UK001 did not use a triage system with the emergency and very urgent triage category.

**S14 Fig. Observed and predicted in % for the number of children for triage categories attending the emergency department for individual sites**

**S15 Fig. Percentage of children admitted to hospital**

Legend:

Percentages of total ED attendances (left) and absolute numbers (right) of children admitted to hospital (top) and pediatric intensive care units (bottom); comparing the 28-day standardized numbers for the months of January – April for 2018 vs. 2019 vs. 2020.

**S16 Fig. The 28-day mean number of selected clinical diagnoses in the emergency department for the period January – April over a three-year period, for high prevalence countries.**

Legend:

Percentages of total ED attendances (left) and absolute numbers (right) of children with diagnosis of a) tonsillitis, b) otitis media, c) lower respiratory tract infections (LRTI), d) gastro-intestinal (GI) infections, e) appendicitis, f) testicular torsion, g) intussusception, h) mental health issues, i) diabetic keto-acidosis, j) radius fracture, k) minor head injury; comparing the 28-day standardized numbers for the months of January – April for 2018 vs. 2019 vs. 2020, shown for countries with of a cumulative 14 day rate of new SARS-CoV-2 cases per 100.000 of 80 or more.

**S17 Fig. 14-day cumulative SARS-CoV-2 case rates for different UK regions**

Legend: the 14-day cumulative SARS-CoV-2 case rate in five different regions in the UK, corresponding with each of the five UK study sites, on May 1^st^ 2020. Source: https://coronavirus.data.gov.uk/details/download

